# Long-read genome sequencing increases genomic yield in congenital heart disease

**DOI:** 10.1101/2025.05.14.25327523

**Authors:** Robert Lesurf, Anjali Jain, Nour Hanafi, Aleksandra Mitina, Yue Yin, Venkat A Kolla, Jade Bouwmeester, Tanya Papaz, Erwin Oechslin, Ryan K C Yuen, Seema Mital

## Abstract

Congenital heart disease (CHD) is the most common birth defect. We performed Illumina short-read genome sequencing (GS) of 1,101 probands, which identified a genetic cause in 16% of cases. We performed PacBio long-read GS in 43 genotype-elusive patients. Paired analysis revealed higher detection with long-read GS of single nucleotide variants, deletions, duplications, and insertions as well as fewer false-positives for indels and inversions. Long-read GS had higher coverage of nine CHD genes, but a low sequencing depth (<10×) in intragenic regions of four of these genes. Long-read GS was better able to resolve complex structural variants and the size of large repeat expansions in 59 known disease-causing regions. This included a complex *de novo* structural variant upstream of *ZEB2* that was only resolved with long-read GS in a patient with extra-cardiac phenotype overlapping Mowat-Wilson syndrome. Long-read GS may provide an option in CHD patients who remain genotype-elusive on short-read GS.

## INTRODUCTION

Congenital heart disease (CHD) is the most common birth defect, occurring in approximately 9 out of every 1000 live births^1^. CHD can consist of singular anomalies, such as septal defects, but also complex anomalies that affect several cardiac structures. Conventional genetic testing identifies a genetic cause of CHD in less than 30% of ‘syndromic’ cases and less than 10% in isolated or ‘non-syndromic’ CHD^2,3^. About 12% of individuals with CHD are found to have chromosomal aneuploidy, and 10-15% harbor a causal microdeletion^4^. Other genetic causes include structural and sequence-level variants that disrupt genes involved in cardiac development.

Short-read genome sequencing (GS) has greatly improved our understanding of the genetic etiology of CHD, revealing new candidate genes and variant types. However, the etiology behind more than half of CHD cases remains unknown. Some of these may be due to environmental causes^5^, and others may be due to genetic variants that are either not captured with current sequencing technologies or where there is insufficient evidence to determine pathogenicity. This includes so-called ‘dark’ regions of the genome, that cannot be accurately assembled or aligned using standard short-read sequencing technologies^6^. Moreover, while short-read GS captures most major variant types, it is often unable to capture regions of the genome that are repetitive or have pseudogenes, and is also ineffective at resolving large structural variants including tandem repeat expansions accurately. The latter have been implicated in neurodevelopmental, neuropsychiatric disorders, cancer^7–11^, and more recently, in cardiomyopathy^12^, but have not been evaluated in CHD. Long-read GS has the potential to overcome these limitations^13^. Further, long-read GS is able to phase variants in the same gene, due to longer read lengths, enhancing our ability to identify pathogenic recessive variants without more costly trio sequencing.

The goal of our study was to compare the genome-wide variant yield, and to specifically compare variants in CHD genes, through paired short-read and long-read GS of CHD patients.

## RESULTS

### Study cohort

We performed short-read GS (Illumina HiSeq X or NovaSeq) on 1,101 CHD probands with two common forms of complex CHD – tetralogy of Fallot (TOF, n=875) and D-transposition of the great arteries (TGA, n=226). Cases were selected from patients enrolled in our Ontario province-wide multi-centre Heart Centre Biobank Registry at the Hospital for Sick Children (Toronto, Canada), the Kids Heart BioBank at the Heart Centre for Children, The Children’s Hospital at Westmead (Sydney, Australia), and the CONCOR registry at the Amsterdam Medical Center (Netherlands)^14^. Probands with a clinically and/or genetically confirmed syndrome were excluded. Previously published analysis of these samples identified 4% patients harboring pathogenic/likely pathogenic disease-associated protein-coding variants, and an additional 1% with canonical and 11% harboring high-confidence non-canonical splice-disrupting variants in CHD genes^14^ (**Table 1**). Probands were deemed genotype-elusive if no pathogenic or likely pathogenic single nucleotide variants (SNVs), insertion-deletions (indels) or copy number variants (CNVs) were identified by clinical genetic testing and/or on short-read GS in Tier 1 CHD genes, which included 99 genes defined as having moderate, strong, or definitive associations with CHD according to Clinical Genome Resource (ClinGen) criteria (17 isolated CHD and 82 syndrome-associated CHD genes)^15^. Variant pathogenicity was determined using the American College of Medical Genetics and Association for Molecular Pathology (ACMG/AMP) guidelines^16,17^. Of 1,052 genotype-elusive patients, 43 underwent additional long-read GS using PacBio. The latter included patients enrolled in the Heart Centre Biobank Registry who had blood-derived DNA available and met one or more of the following criteria: parent-child trio DNA available, presence of extra-cardiac features not consistent with a defined syndrome, and/or availability of myocardial RNA for variant validation.

The study was approved by local Research Ethics Boards, written informed consent was obtained from all patients, parents or legal guardians, and study protocols adhered to the Declaration of Helsinki.

### Sequencing quality between short-read and long-read GS

To first determine whether DNA extractions intended for short-read GS were also viable for long-read GS, we compared the DNA fragment sizes in previously extracted and stored DNA samples that used a conventional protocol versus long fragment DNA extracted from fresh whole blood using a targeted protocol. Targeted long-fragment DNA extraction from a fresh blood sample yielded 99.8% fragments ≥ 20 kb in length, 95.8% fragments ≥ 50 kb in length, and a DNA integrity number (DIN) of 9.8. DNA previously extracted and stored from whole blood samples using a conventional extraction protocol suitable for a wide range of downstream applications yielded 80%-93% fragments ≥ 20 kb in length, 67%-87% fragments ≥ 50 kb in length, and DINs ranging from 8.2-9.4 (**Figure 1a**, **Table 2**). DNA samples from 43 unrelated probands (36 TOF, 7 TGA) having at least 70% fragments ≥ 20 kb in length and a DIN ≥ 8 were selected for long-read GS using PacBio Sequel IIe platform (**Figure 1b**, **Table 1).** There was no significant association between the initial fragment sizes or DINs and the resulting mean long-read sequenced read length (p-value > 0.05) (**Figure 1c-e**). Mean genome-wide sequencing depth of short-read GS was 31.8× (range 24× to 46×) while that of long-read GS was 17.9× (range 11× to 22×) (**Supplemental Figure S1a**). For the purposes of this study, we focused all subsequent analyses on the 43 probands with paired short-read and long-read GS.

**Figure 1.**
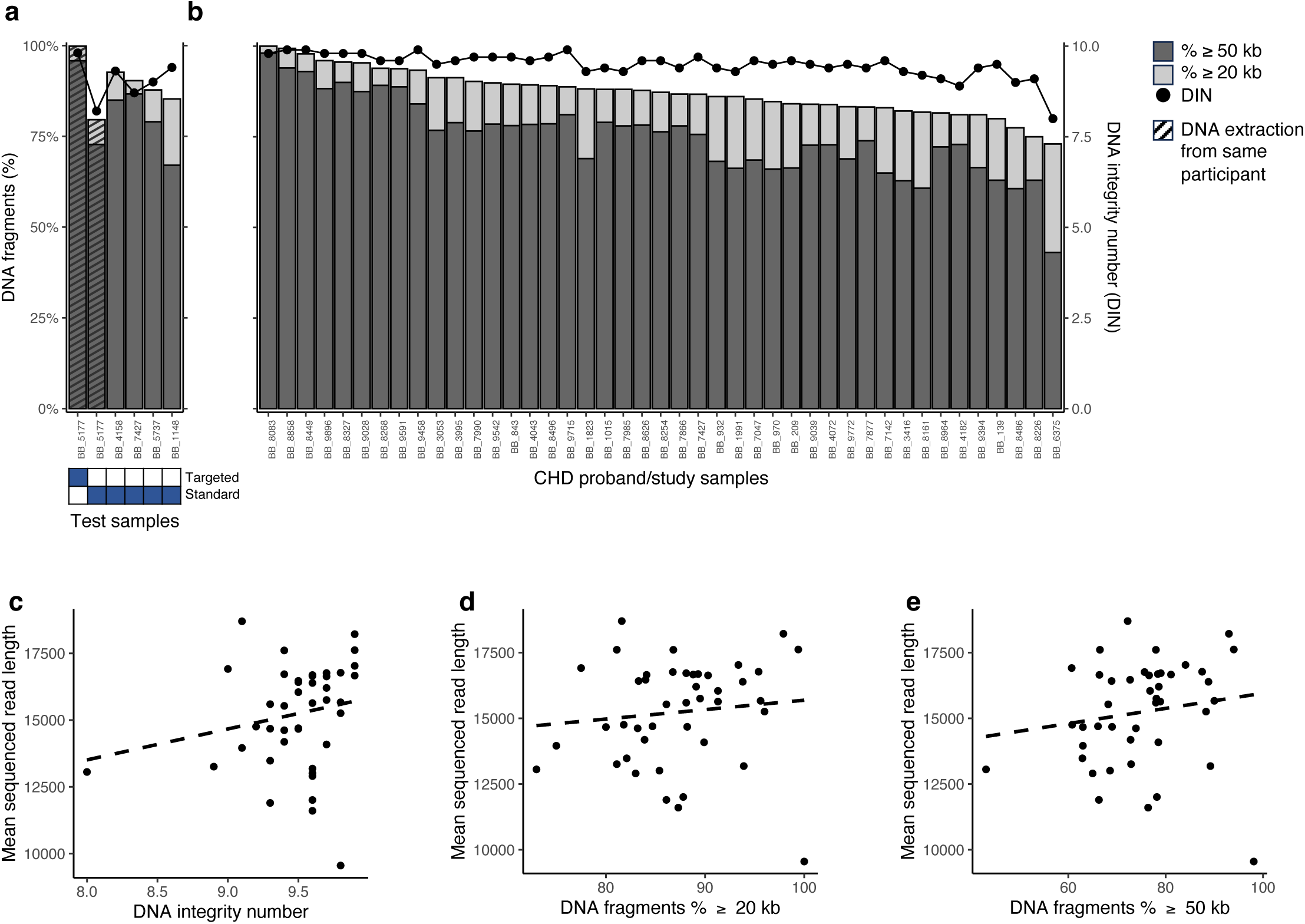
Long-fragment DNA extraction quality control metrics. **(a)** Percentage of DNA fragments greater than or equal to 20 kb (light grey) and 50 kb (dark grey) in length, and DNA integrity number (DIN) for five previously extracted DNA samples using a standard protocol and one DNA extraction using a targeted high molecular weight kit specifically designed for long-read sequencing. One participant’s whole blood was used for both methods of DNA extractions, as denoted by hashed lines. **(b)** Percentage of DNA fragments greater than or equal to 20 kb and 50 kb in length, and DNA integrity number (DIN) for 43 previously extracted and frozen DNA samples from unrelated CHD probands that met quality metrics were subsequently used for long-read sequencing. **(c)** Association of mean sequenced read length with DIN (n = 43, Pearson r = 0.20). **(d)** Association of mean sequenced read length with percentage of extracted DNA fragments ≥ 20 kb (n = 43, Pearson r = 0.11). **(e)** Association of mean sequenced read length with percentage of extracted DNA fragments ≥ 50 kb (n = 43, Pearson r = 0.16). There was no significant association between mean long-read sequenced read length and DIN or fragment size (p-value > 0.05). DNA, deoxyribonucleic acid; DIN, DNA integrity number; CHD, congenital heart disease

There were no sex mismatches between reported and predicted sex in short-read and long-read GS (**Supplemental Figure S1b**). Genetic relationships analyzed among the short-read and long-read GS data confirmed low relatedness scores between all probands (**Supplemental Figure S1c**). Genomically-predicted ancestry of all probands matched with self-reported ancestry (**Supplemental Figure S1d**). The majority of the cohort had European ancestry (n=35; 81%), followed by South Asian (n=4; 9%), East Asian (n=2; 5%), and admixed (n=2; 5%) **(Table 1)**.

### Genome-wide and CHD gene-specific variant calls in short-read and long-read GS

*Small variants*: We compared the yield of single nucleotide variations (SNVs) and insertion-deletions (indels) that were called genome-wide, as well as variants that overlapped Tier 1 CHD genes (**Table 3**).

Long-read GS captured a higher number of SNVs than short-read GS, both genome-wide (1.01-fold) and in Tier 1 CHD genes (1.01-fold) (paired Wilcoxon rank-sum test, p-value < 0.05) (**Figure 2a, Supplemental Figure S2a**). The opposite trend was observed for indels, with a higher number of indel calls with short-read GS compared to long-read GS, both genome-wide (1.01-fold) and in Tier 1 CHD genes (>1.00-fold) (**Figure 2b, Supplemental Figure S2b**). We then compared Jaccard index in variant calls, defined as the intersect of variants called in both short-read and long-read GS divided by the union of all calls. We observed a high average Jaccard index for SNVs at 0.88 genome-wide and 0.94 in Tier 1 CHD genes. For indels, the average Jaccard index was lower between the two sequencing technologies, with 0.73 genome-wide and 0.72 in CHD genes. On average, 69.7% and 70.5% of indel calls in short-read and long-read, respectively, fell in low complexity regions, even though these regions make up only 2% of the genome^18^. Previous reports have found that long-read sequencing has higher accuracy for calling indels, particularly in repetitive regions of the genome^19,20^, suggesting that the indels only called in our short-read GS samples were more likely to be false positives.

**Figure 2.**
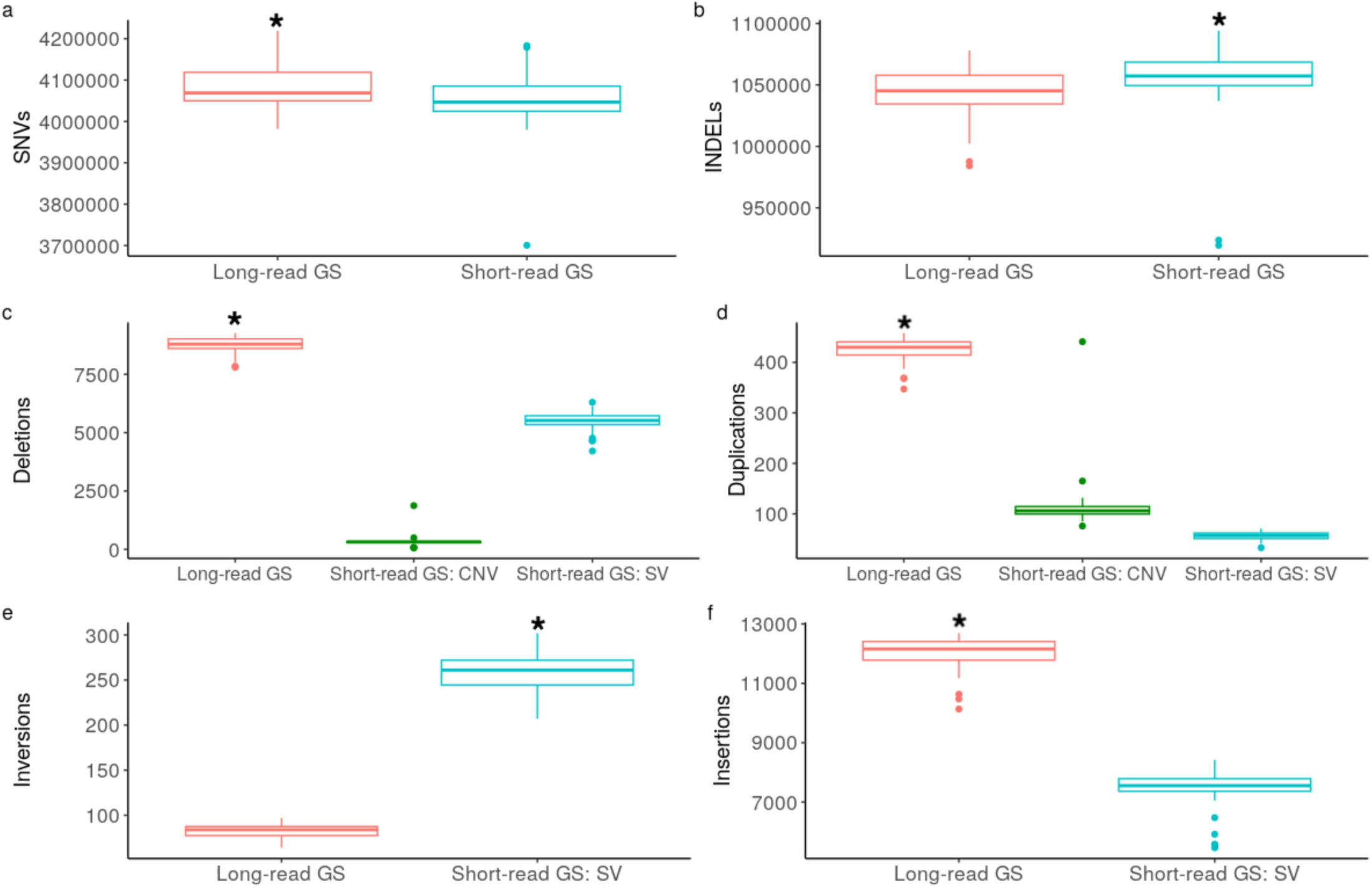
Genome-wide variant yield by long-read and short-read genome sequencing. Box plots comparing variants called in long-read GS and short-read GS, including **(a)** SNVs, **(b)** Indels, **(c)** Deletions, **(d)** Duplications, **(e)** Inversions, and **(f)** Insertions. Genome-wide, long-read GS called more SNVs (1.008-fold; nominal p-value = 3.57×10^-6^), deletions (1.604-fold; nominal p-value =2.0×10^-25^), duplications (7.734-fold; nominal p-value =5.2×10^-25^), and insertions (1.634-fold; nominal p-value = 1.5×10^-15^) than short-read GS but fewer indels (0.99-fold; nominal p-value 3.3×10^-6^) and inversions (0.325-fold; nominal p-value = 1.4×10^-15^). Short-read GS: CNV designates structural variants called by copy number variant callers, while Short-read GS: SV designates structural variants called by structural variant callers. Asterisk denotes significant differences (nominal p-value < 0.05). SNV, single nucleotide variants; Indels, small insertion-deletions; CNV, copy number variants; SV, structural variants

*Structural variants*: We next compared the number of structural variants (SVs), including copy number variants (CNVs), called by each platform. Long-read GS called 1.60-fold more deletions, 7.73-fold duplications, and 1.64-fold insertions, but 3.07-fold fewer inversions genome-wide and in Tier 1 CHD genes, likely attributable to fewer false positives in the long-read pipeline^19^. (**Figure 2c-f, Supplemental Figure S2c-f**).

To compare the Jaccard index of SVs genome-wide between long-read and short-read methodologies, we intersected long-read SVs with those from short-read. As the exact breakpoints of SVs are difficult for callers to accurately define, a SV was defined as intersecting if it had a 50% reciprocal overlap between calls made in long-read and short-read GS data. The Jaccard index was highest for deletions at 0.46, followed by insertions at 0.32, inversions at 0.17, and lowest for duplications at 0.04 (**Figure 3a**, **Table 4**). These observations are consistent with previous reports that deletions are relatively easy to identify using short-read sequencing due to spanning reads^20^, while insertions are more challenging due to difficulties in mapping a novel insertion site not present in the reference genome, or in trying to map an insertion that is duplicated genome-wide and thus ambiguous^21^. Duplications are also challenging to detect using short-read sequencing, particularly in segmental duplication sites that have increased copy number gains genome-wide, due to a diffused depth signal caused by ambiguous mapping^22^. To investigate whether these disagreements were driven by SV size, we compared the size distributions of the long-read SVs to the short-read SVs, and plotted the genome-wide density distributions of the SV lengths stratified by SV type. Visual analysis showed that the peaks and valleys of the deletion size distributions were generally similar, further indicating agreement in deletion calls between the two technologies (**Figure 3b**). However, the median length of duplications and inversions was significantly larger in short-read GS calls (**Figure 3c-d**, **Table 4**). This is consistent with a previous study that identified an overrepresentation of very large SV calls in short-read data that are false positives^23^.

**Figure 3.**
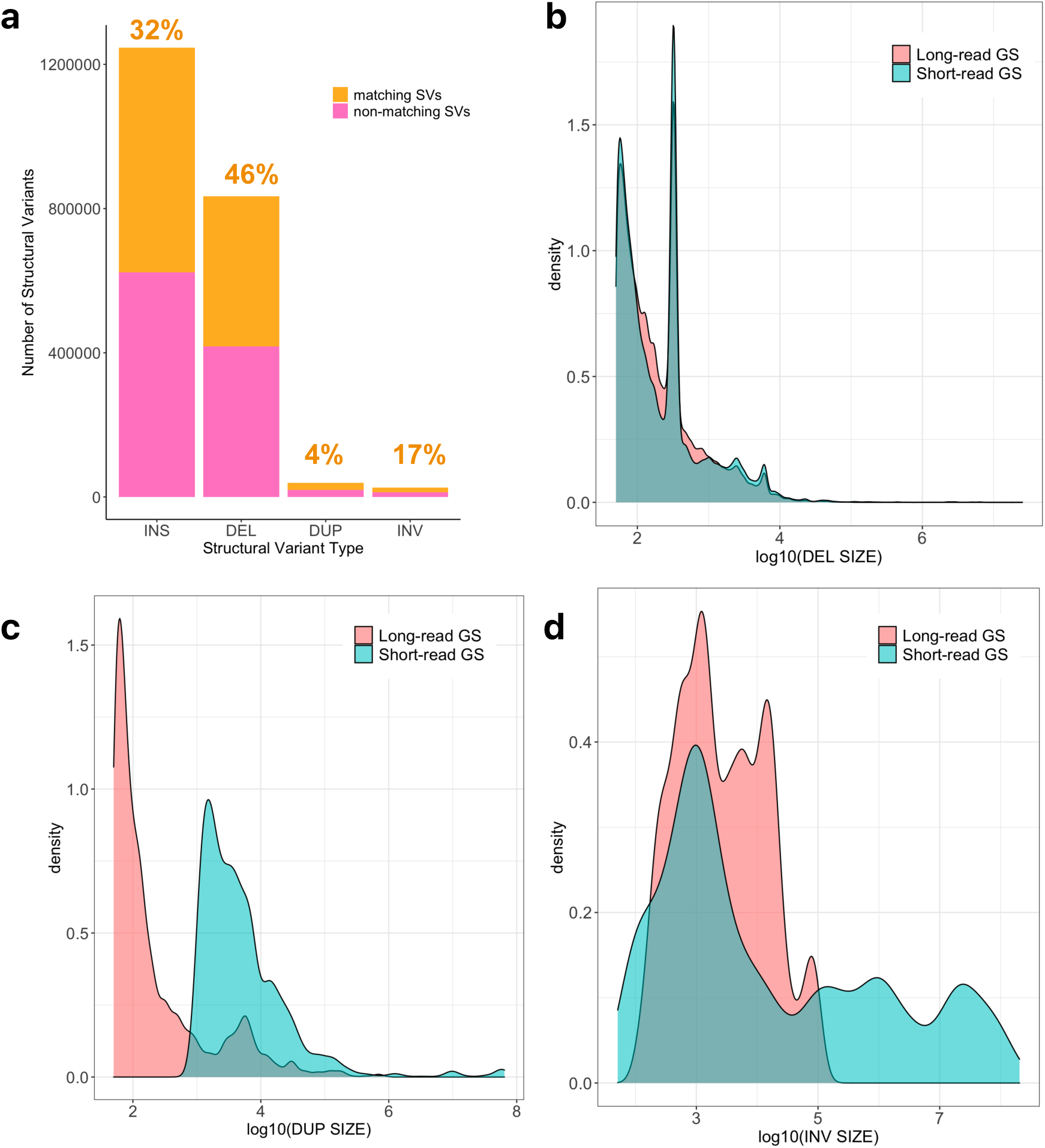
Structural variant calls by long-read and short-read genome sequencing. (**a**) Proportion of matching structural variants called in long-read and short-read GS data. Text labels indicate the proportion of shared variants between the two technologies, as defined by a minimal reciprocal overlap of 50. (**b-d**) Density distribution plots of deletions, duplications and inversions larger than 50 bp, by long-read and short-read GS. Deletion calls were similar across nearly all sizes for both GS methods, while long-read GS tended to call smaller length duplications and inversions. GS, genome sequencing; SV, structural variant; INS, insertion; DEL, deletion; DUP, duplication; INV, inversion

We further investigated whether different types of insertion calls were discordant between the long-read and short-read GS data. As only the standard long-read SV calling workflow provided annotations for the types of insertions called, we calculated the proportion of long-read insertions that were also called in short-read data to estimate concordance for calling each insertion type.

Alu and L1 insertion long-read calls had a consistently high concordance across samples (**Supplemental Figure S3**, **Table 4**). SINE-VNTR-Alu retrotransposon (SVA) insertions also had a high median but variable concordance across samples, likely attributable to the small number of SVA calls made in each sample. Conversely, a much lower concordance was observed for tandem repeat insertions between long-read and short-read samples. To further explore these findings, we next employed additional tools designed to more accurately estimate tandem repeat sizes in long-read and short-read data.

*Tandem repeat expansions*: To more accurately compare the ability of long-read and short-read GS to resolve tandem repeat lengths, we derived repeat estimates using different computational tools designed for use with each type of technology. For long-read GS, we applied the Tandem Repeat Genotyping Tool^24^ to all probands in 59 known disease-causing tandem repeat regions, and similarly used ExpansionHunter^25^ for short-read sequencing data from these patients applied to the same coordinates and repeat motif sequence for the same regions. Nine of these 59 regions were located in or adjacent to Tier 1 CHD or other cardiac relevant genes (**Table 5**).

A majority of the 59 tandem repeat regions (86%) had a Pearson’s correlation coefficient > 0.5 between long-read and short-read GS estimates for both larger and smaller sized alleles, suggesting that tandem repeat lengths can be accurately sized using either sequencing approach for most regions (**Figure 4**, **Table 5**). However, for two regions, in *HRAS* (a Tier 1 CHD gene) and *XYLT1*, long-read GS provided a more reliable and precise estimation of repeat size (**Supplemental Figures S4-S5**). The accurate sizing of these regions may be challenging with short-read GS, primarily due to their large repeat size (∼30-unit GGCGTCCCCTGGAGAGAAGGGCGAGTGT-repeat in case of *HRAS*, and a ∼100-unit GCC-repeat in case of *XYLT1*).

**Figure 4.**
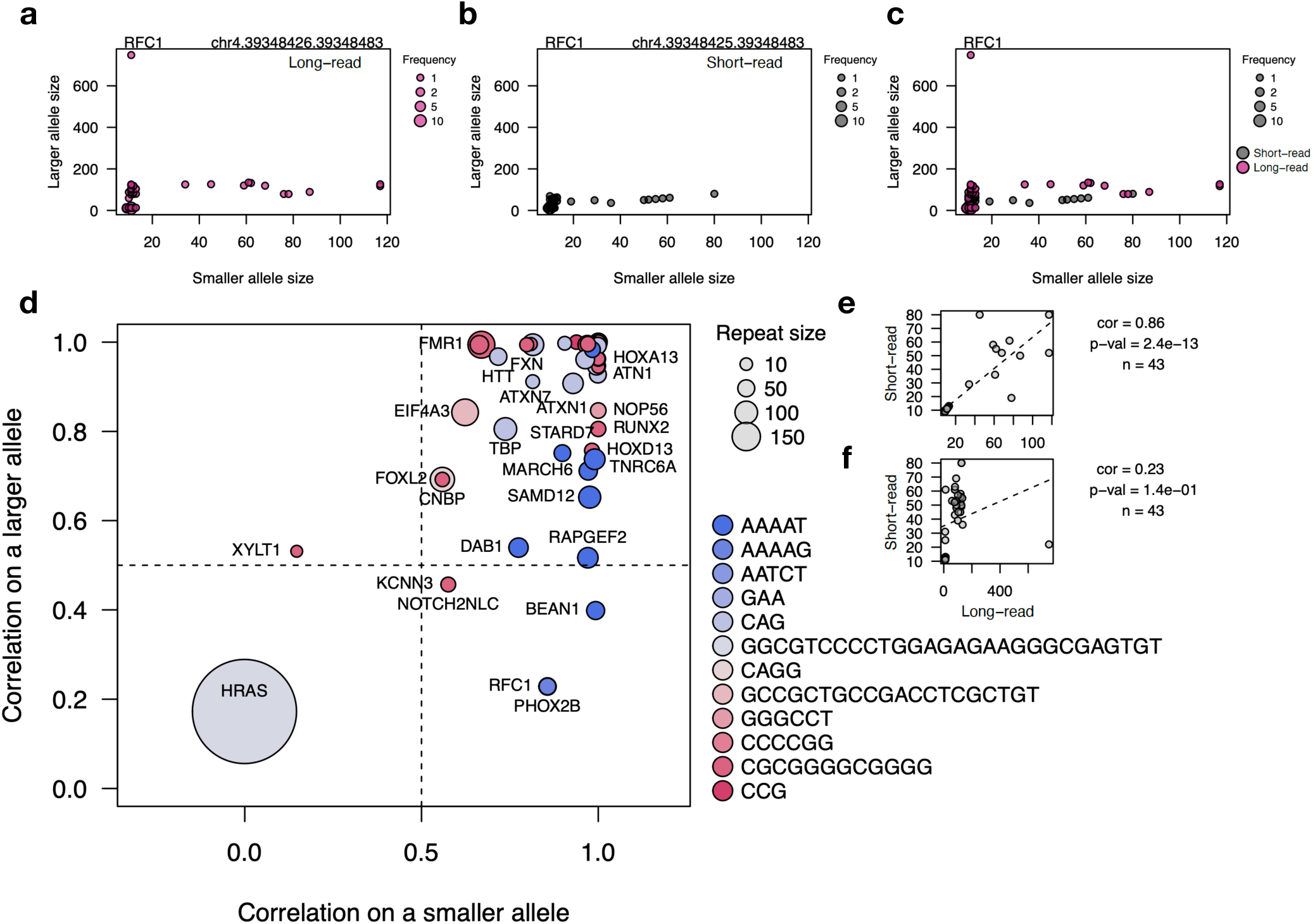
Tandem repeat sizing by short-read and long-read sequencing. AAAAG-repeat near *RFC1* gene detected using **(a)** long-read sequencing (pink), **(b)** short-read sequencing (grey, and **(c)** comparison of the two platforms. X-axis represents repeat size on a smaller allele, y-axis represents size on a larger allele. Each dot represents a genotype; the size of the dot represents the number of samples with this genotype. Smaller and larger allele size estimates are similar for most participants across either platform, but in 3 participants, only long-read sequencing detected large outlier allele sizes. **(d)** Correlation between the size estimated by long-read and short-read sequencing on a smaller allele (x-axis) and a larger allele (y-axis) across all 59 repeat regions (**Table 5**). Each dot represents one repeat region; the size of the dot corresponds to the repeat size in a reference genome; color corresponds to the repeat motif. Most repeats near genes had high correlation for both the smaller and larger allele, but we observed regions with correlations < 0.5 on the smaller allele and/or larger allele, indicating poor agreement between short-read and long-read GS. Size estimates for the 28-base pair repeat near *HRAS* – the longest repeat studied – had low correlation for both smaller and larger alleles, indicating that even a small number of repeats could not be resolved by short-read sequencing. **e,f.** Comparison of **(e)** smaller allele size, and **(f)** larger allele size in short-read (x-axis) vs long-read (y-axis) sequencing data for the *RFC1* gene. Pearson’s correlation coefficient and p-value are labeled beside each plot. Estimates for smaller alleles were significantly correlated between the two platforms, suggesting that both short-read and long-read sequencing generally provided accurate estimates for smaller alleles. However, long-read sequencing provided more accurate estimates for larger alleles, as indicated by the non-significant correlation between the two platforms.

In another set of regions, short-read GS was able to estimate repeat length accurately on the smaller allele, but not on the larger allele (i.e., poor correlation with long-read GS estimates).

These regions were in *BEAN1*, *DAB1*, *MARCH6*, *PHOX2B*, *RAPGEF2*, *RFC1*, *SAMD12*, and *TNRC6A* (**Figure 4d** and **Supplemental Figure S6**). It should be noted, however, that within the regions where the disease-associated motif was different from the reference motif, such as *BEAN1*, *DAB1*, *RFC1* and others (**Table 5**), the larger repeat contained the reference motif only and was therefore not considered pathogenic.

### Sequencing depth of CHD genes in long-read and short-read GS

We next systematically investigated the sequencing depth of 147 CHD genes which included 99 Tier 1 CHD genes, and 48 additional cardiac relevant genes from the Twist Alliance Dark Genes Panel^26^ that reside in the so-called ‘dark’ regions of the genome, indicating that they are challenging to sequence and/or align with short-read GS^6^. Of note, eight Tier 1 CHD genes (*ADAMTS10*, *B3GAT3*, *BRAF*, *EHMT1*, *FLT4*, *G6PC3*, *KANSL1*, and *PKD1*) were included in the Twist panel **(Table 3)**.

In our cohort, long-read and short-read samples were sequenced to different genome-wide target sequencing depths of ∼20× and ∼30×, respectively. Therefore, as expected, most individual genes had higher sequencing depths in short-read GS samples, except for *CFC1*, *CFC1B*, *H19*, and *SMN1* (**Figure 5a**, **Table 6**). To enable us to identify where each technology offers specific relative advantages and disadvantages, we further calculated normalized sequencing depths (i.e., gene sequencing depth divided by the genome-wide sequencing depth) in all samples for the two technologies for the 147 genes of interest (**Table 6**). A normalized sequencing depth value below 1 indicates that the gene had fewer aligned reads than the average observed depth across the genome, and a value greater than 1 indicates that the gene had a higher sequencing depth than the average observed depth across the genome. 121 of 147 genes had a statistically significant difference in the normalized sequencing depth between long-read and short-read (false discovery rate, FDR < 0.05), while the remaining 26 genes had no significant difference. Of the 121 genes with a significant difference, 112 genes had a higher median normalized sequencing depth in short-read and nine genes had a higher median normalized sequencing depth in long-read **(Figure 5b**, **Table 6).**

**Figure 5.**
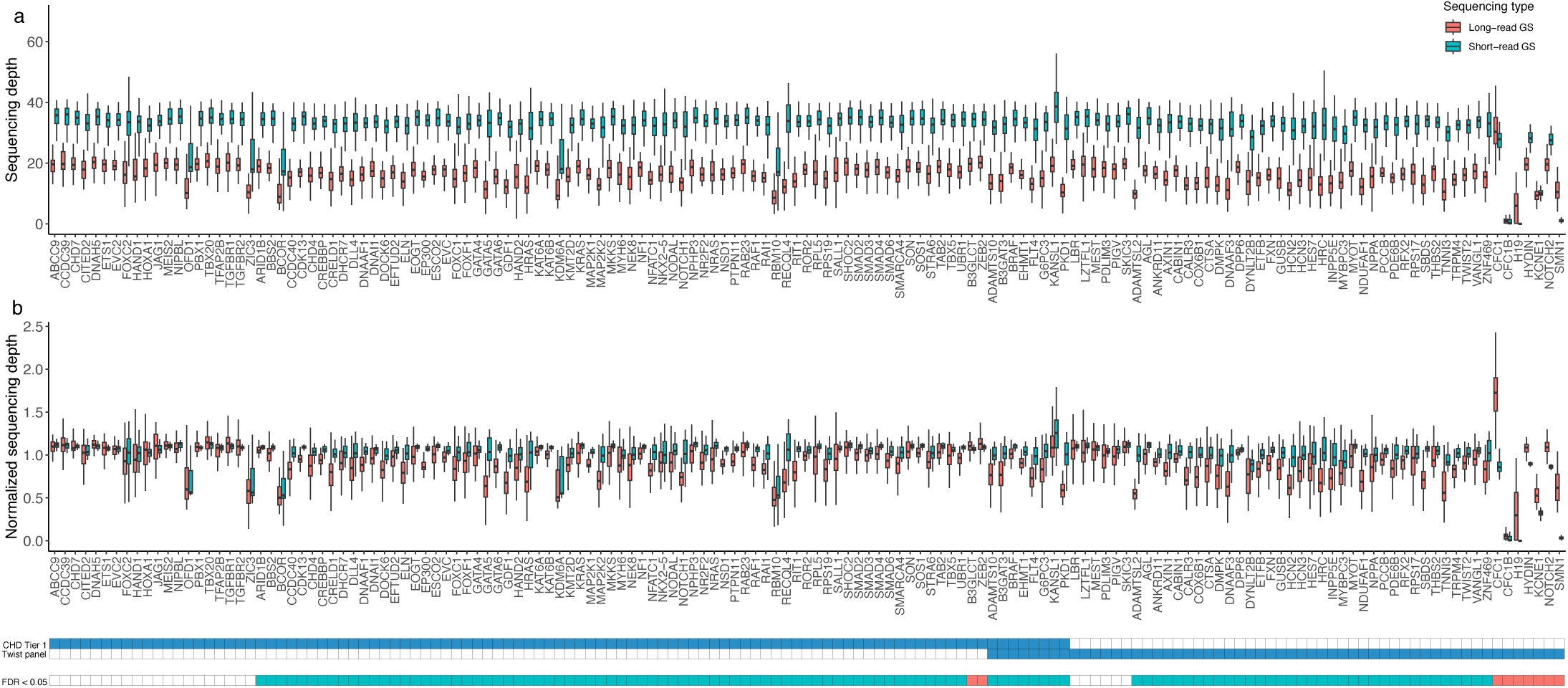
Sequencing depth comparison of CHD genes. **(a)** Sequencing depth and **(b)** normalized sequencing depth (i.e., gene sequencing depth divided by the genome-wide sequencing depth of a sample) of 147 CHD genes (99 Tier 1 genes and 48 cardiac relevant genes from the Twist panel) in long-read GS (red) and short-read GS (blue). 121 genes had a statistically significant difference (FDR < 0.05) in the normalized sequencing depth between the two technologies whereas, 26 genes had no significant difference (white in the bottom matrix). Out of the 121 genes with a significant difference, 112 genes had a higher median normalized sequencing depth in short-read (blue in the bottom matrix) and, 9 genes had a higher median normalized sequencing depth in long-read (red in bottom matrix). CHD, congenital heart disease; GS, genome sequencing, FDR, false discovery rate

Among the nine genes that had a significantly higher normalized sequencing depth in long-read GS samples, (**Figure 6**), two genes (*B3GLCT* and *ZEB2*) still had consistently good gene-wide normalized sequencing depths in short-read data (**Figure 6a-b**), while three genes (*CFC1*, *HYDIN* and *NOTCH2*) had lower normalized sequencing depth with short-read compared to long-read GS (**Figure 6c-e**). An intragenic analysis revealed that the sequencing depth of these three genes was non-uniform in short-read GS compared to long-read GS, which provided uniform coverage across all exonic and intronic regions of these genes (**Figure 7a-c**, **Table 7**). The lower short-read sequencing depths observed in our cohort were consistent with those reported in gnomAD v3 short-read GS control data (**Figure 7a-c)**. Moreover, we visually noted that the entirety of the *CFC1* gene and ∼3.5kb of the *HYDIN* gene falls into “LowMap+SegDup” regions (i.e., low mappability and segmental duplication) in the Genome in a Bottle track^27^ in the UCSC genome browser and similarly noted low mappability in the above mentioned regions of *NOTCH2* using the multi-read mappability with 24-mers (M24) track by Umap^28^ in the UCSC genome browser. Finally, despite having a significantly higher median normalized sequencing depth in long-read GS compared to short-read GS, the normalized sequencing depth for the remaining four genes (*CFC1B*, *H19*, *KCNE1*, and *SMN1*) remained below 1 for most samples indicating that even long-read GS was not able to fully resolve these genes at the target sequencing depth we used (**Figure 6 f-i**). Specifically, a majority of the long-read GS samples had a raw sequencing depth of < 10× for all exons and introns of *CFC1B* and *H19*, exon 3-4 & intron 3 of *KCNE1* and exon 1 & intron 1 of *SMN1* **(Figure 7d-g)**. Therefore, sequencing depth may need to be higher when specifically targeting these genes even with long-read GS.

**Figure 6.**
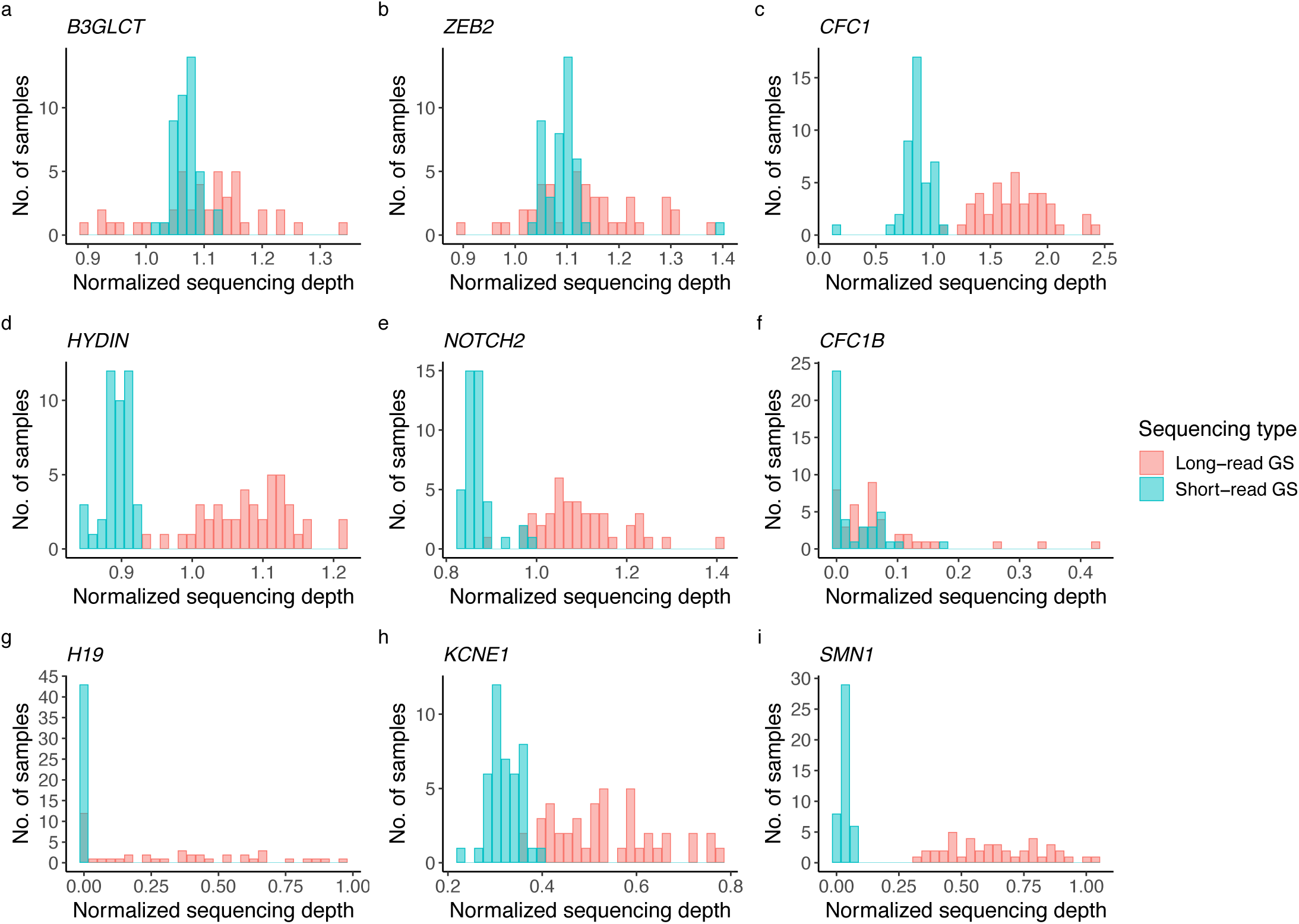
Distribution of normalized sequencing depth in genes with a higher depth in long-read GS. Histograms comparing the normalized sequencing depth (i.e., gene sequencing depth divided by the genome-wide sequencing depth for each sample) distribution across nine genes with a significantly (q-value < 0.05) higher median sequencing depth in long-read GS (red) than short-read GS (blue). **(a)** *B3GLCT*. **(b)** *ZEB2. **(c***) *CFC1*. **(d)** *HYDIN*. **(e)** *NOTCH2*. **(f)** *CFC1B*. **(g)** *H19.* **(h)** *KCNE1.* **(i)** *SMN1*. For *B3GLCT* and *ZEB2*, long-read GS had a higher normalized sequencing depth, and short-read GS had a lower but adequate median normalized depth of 1.09 and 1.07 respectively. For *CFC1*, *HYDIN*, and *NOTCH2,* there was a higher depth in long-read than short-read GS i.e. median greater than 1. For *CFC1B*, *H19*, *KCNE1* and *SMN1*, although normalized sequencing depth was higher for long-read compared to short-read, the normalized sequencing depth was still below 1 for all samples, indicating suboptimal coverage for these genes even with long-read GS. GS, genome sequencing

To investigate whether these differences in sequencing depth affected variant calling, we calculated the median Jaccard indices for the individual introns and exons of each gene included in **Figure 7**. This comparison was limited because of the small number of intragenic variants called by either sequencing technology, but suggested that a very low sequencing depth resulted in fewer variant calls. For example, in *NOTCH2*, two introns (1 & 4) had a Jaccard index < 0.8, which both also had a significantly lower normalized sequencing depth in short-read data (**Table 7**). Similarly, intron 2 of *KCNE1* and intron 7 of *SMN1* had low agreements of variant calls (median Jaccard index = 0.7 and 0, respectively), reflecting the significant difference in normalized sequencing depth; no variants were called in other regions of these two genes.

**Figure 7.**
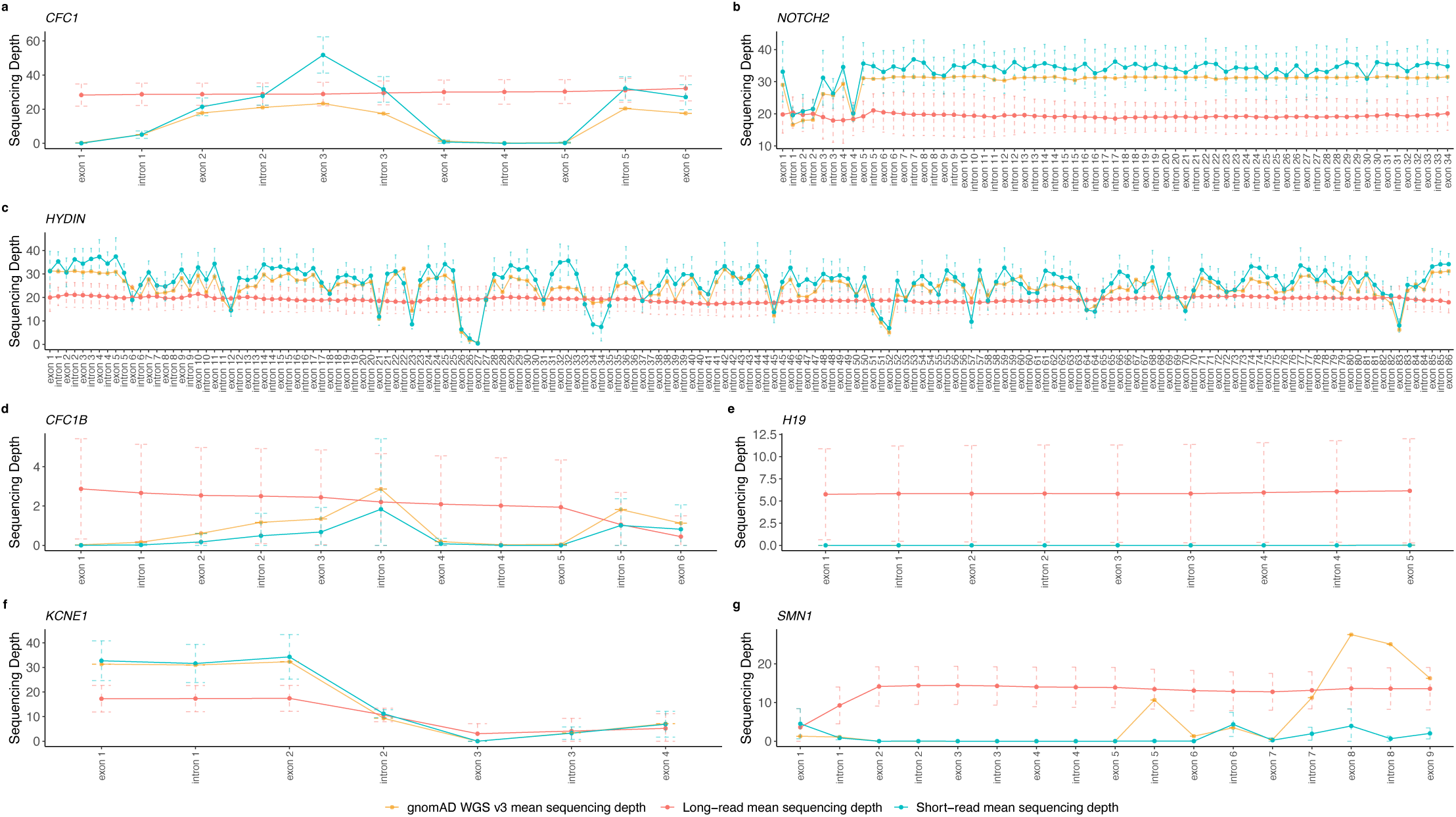
Intragenic sequencing depth comparison for *CFC1*, *NOTCH2*, *HYDIN*, *CFC1B*, *H19*, *KCNE1* and *SMN1*. Mean sequencing depth (y-axis) for every exon and intron of (**a**) *CFC1*, (**b**) *NOTCH2*, (**c**) *HYDIN*, (d) *CFC1B*, (e) *H19*, (f) *KCNE1*, and (**g**) *SMN1* in the long-read (red), short-read (blue), and gnomAD v3 (yellow) sequencing cohorts. The dashed bars represent the error bars calculated as the mean sequencing depth ± standard deviation. Short-read sequencing depth in our study cohort was similar to that in the gnomAD short-read GS samples. For *CFC1* and *HYDIN*, long-read GS had uniform sequencing depth across all gene regions whereas the sequencing depth was non-uniform with short-read sequencing. for *NOTCH2*, short-read GS has a higher sequencing depth across all gene regions, however, there are drops in sequencing depth in exon 2 & introns 1-4 whereas long-read sequencing displayed a uniform sequencing depth. For *CFC1B*, while the sequencing depth in long-read GS was consistently higher than short-read GS, the mean sequencing depth in long-read GS was low i.e. < 5× across the entire cohort. Similarly, for H19, most samples had a sequencing depth < 10× across all exons and introns in long-read GS. For *KCNE1* both long-read and short-read GS struggled to sequence the intergenic region of *KCNE1* encompassing exon 3, intron 3 and exon 4. Lastly, for *SMN1*, long-read GS had consistently higher sequencing depth than short read GS for all exons and introns except exon 1 and intron 1 where the sequencing depth was < 10×. GS, genome sequencing

Conversely, among the intragenic regions of *HYDIN* that had a higher median normalized depth in long read GS, we observed similar variant calls for 19 introns (median Jaccard index > 0.8) and a low agreement for only one intron (median Jaccard index = 0.6) (**Table 7**). There were no variants called in the remaining exons and introns of *HYDIN* that had a higher median normalized depth in long-read GS. This broad agreement between sequencing platforms reflects the visual observation that most intragenic regions of *HYDIN* appeared to have a high enough sequencing depth in both long-read and short-read GS to reliably call variants. Finally, there were no variants called in *CFC1, CFC1B* and *H19*, which prevented a comparison of these remaining genes.

### Identification of putatively pathogenic variants in CHD genes by long-read GS

In this cohort of 43 patients who were genotype-elusive on short-read sequencing, we further searched for putatively pathogenic variants in CHD genes. This included searching for SNVs, indels, CNVs, and SVs that were rare in control cohorts (gnomAD v2 short-read GS and whole exome sequencing data (n=14,891)^29^, gnomAD v3 GS data (n=76,156)^30^, Genomic Answers for Kids (GA4K) long-read GS data (n=960)^31^, and the Database of Genomic Variants short-read GS data (n=14,316)^32^; full details in methods). Analysis of long-read GS did not identify any definitively pathogenic or likely pathogenic variants in Tier 1 CHD genes. However, we did identify putatively damaging variants in CHD genes that were only called in long-read, or in one case, in short-read GS data.

We identified a TOF proband with developmental delay, autism, microcephaly and joint hyperlaxity that was intensively investigated with a chromosomal microarray, multiplex-ligation dependent probe amplification for 22q11 deletion, testing for Angelman syndrome, various metabolic tests, and short-read whole exome sequencing all of which returned negative results. Short-read GS called both a breakend and an insertion at GRCh38 chr2:138333831, an insertion at chr2:144181512, and a breakend at chr2:144181513, encompassing the syndromic CHD gene *KYNU*. Closer inspection of both the short-read and long-read breakends suggested that this variant was actually an insertion of the chr2 region into chr10, with a smaller inversion sharing the right breakend on the alternate allele (**Figure 8**). Long-read sequencing correctly called this variant as a breakend. This variant was absent in the short-read genomes of the parents and thus appeared to be *de novo.* Although the patient did not harbor the complete phenotype of vertebral, cardiac, renal and limb defects syndrome 2 (VCRL2), the autosomal recessive disease associated with *KYNU,* they had several features compatible with Mowat-Wilson syndrome (heart defect, microcephaly, developmental delay), a condition associated with heterozygous disruption of the gene *ZEB2*. Interestingly, *ZEB2* is located 202 kbp downstream of this variant – the translocation of the region to chr10 may therefore have resulted in disruption to *ZEB2* gene regulation. We did not have access to myocardial tissue in this patient to confirm changes in gene expression.

**Figure 8.**
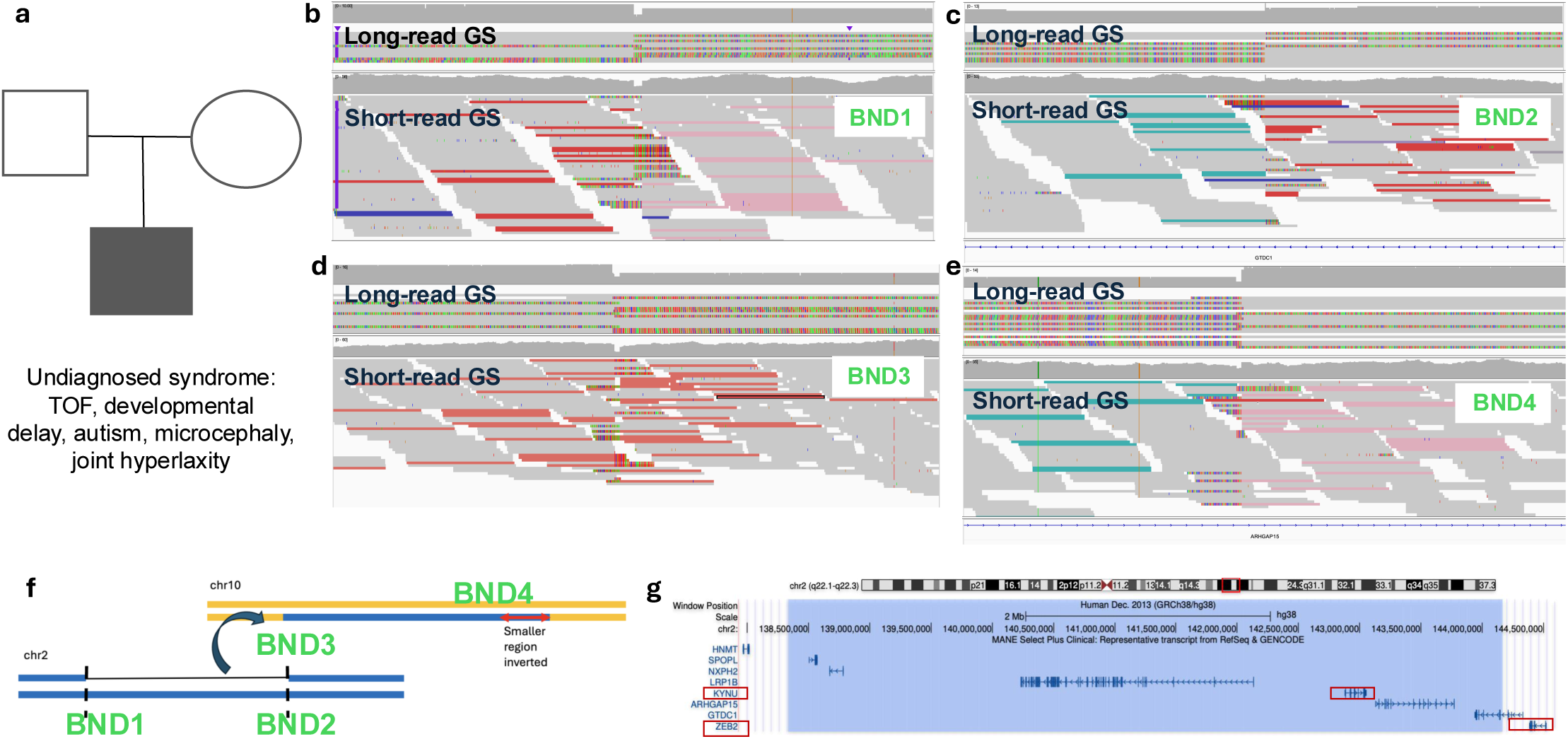
Complex structural variant associated with syndromic CHD. A translocation-inversion event identified in a patient with syndromic CHD. **(a)** Pedigree of affected individual. **(b)** Reads aligned to the left breakend (BND) on chr2. Top panel = long-read GS, bottom panel = short-read GS. Pink reads indicate a mate mapping to chr10. Red reads indicate a larger than expected insert size. Rainbow colored read portions indicate bases that are mismatching relative to the GRCh38 reference, suggesting those sequences derived from elsewhere in the genome. **(c)** Reads aligned to the right BND on chr2. Teal reads indicate the presence of an inversion event. **(d)** Reads aligned to the insertion site on chr10. Light red reads indicate mates mapping to chr2. **(e)** Displays the right BND of the inversion, with pink reads indicating mapping to chr10. **(f)** A proposed schematic of the complex structural variant, indicating that one copy of the locus was translocated into chr10, with a smaller section of the locus inverted. **(g)** Representation of the protein-coding genes within the translocated chr2 region, within the blue box. The cardiac-relevant genes of interest, *KYNU* and *ZEB2* are outlined in red. CHD, congenital heart disease; GS, genome sequencing; TOF, Tetralogy of Fallot, BND, breakend

Another TOF proband was found to harbor compound heterozygous SNVs in the syndromic CHD gene *NPHP4*, which is associated with Senior-Løken syndrome, an autosomal recessive ciliopathy. Long-read sequencing allowed for the phasing of the variants to separate parental alleles and confirmed that the SNVs were in trans **(Figure 9a).** At last follow-up before 5 years of age, there was no evidence of nephronophthisis although these findings may not manifest until later in childhood.

**Figure 9.**
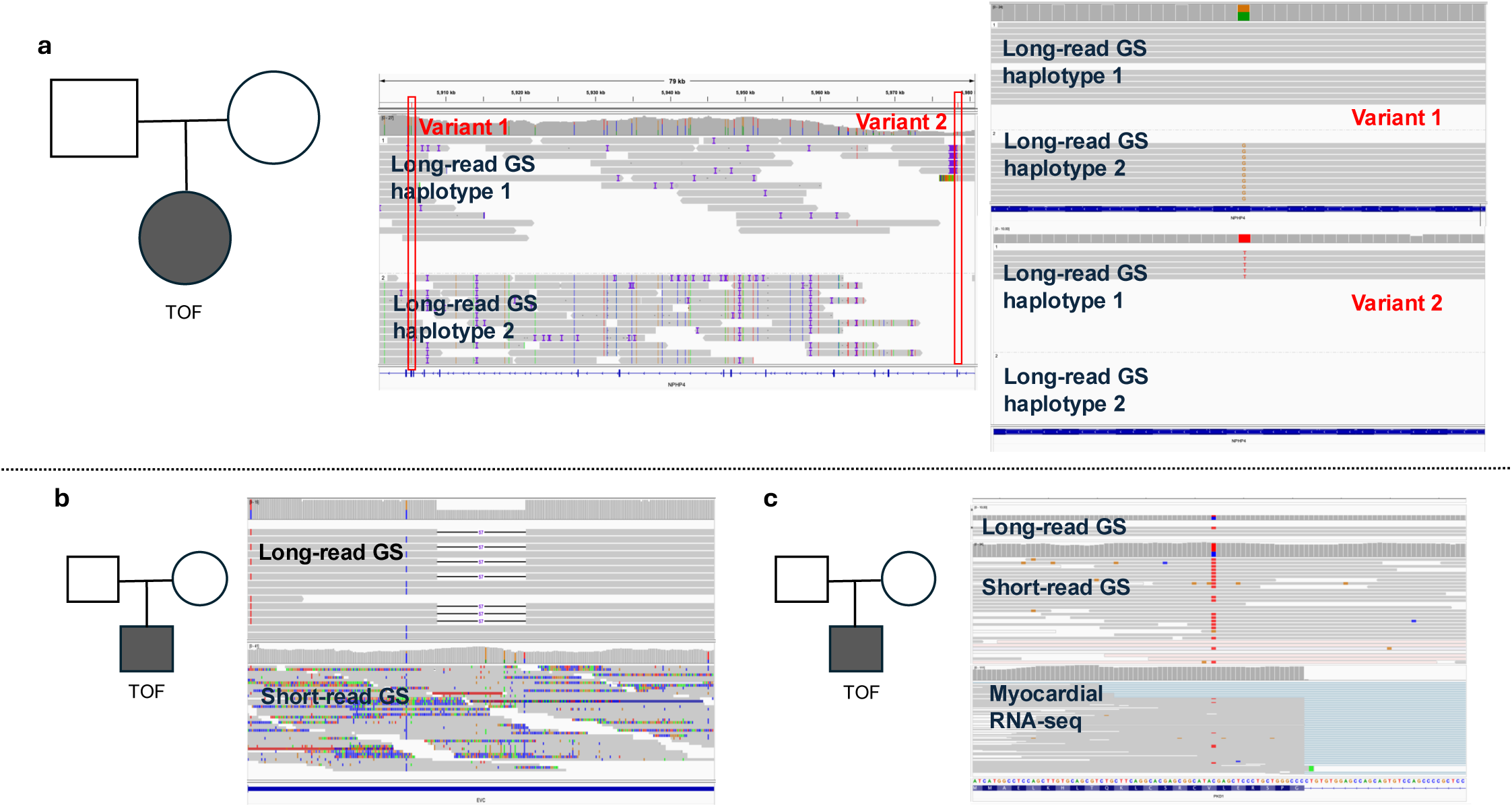
CHD gene high-risk variants. **(a)** A 57-base pair deletion in the syndromic CHD gene *EVC*, identified by long-read (top panel) but not short-read (lower panel). **(b)** Two variants in the syndromic CHD gene *NPHP1*, 72,942 bp apart, were phased as *trans* by long-read GS, but could not be phased by short-read sequencing in the absence of trio sequencing. The panels represent the two different parental haplotypes. **(c)** A SNV in *PDK1* with only one read of support was not called in long-read GS data (upper panel) but was called in short-read GS data (middle panel). The SNV was predicted to create a cryptic splice site, which was validated using short-read myocardial RNA sequencing from patient (bottom panel). CHD, congenital heart disease; TOF, Tetralogy of Fallot; SNV, single nucleotide variant; GS, genome sequencing; RNA-seq, RNA sequencing

A third case involved a TOF proband in whom long-read GS identified a 57 bp deletion in the final exon of *EVC*, a Tier 1 CHD gene associated with Weyers acrofacial dystosis and Ellis-van Creveld syndrome, although the patient did not demonstrate characteristic features associated with this syndrome (**Figure 9b**). Therefore, despite the small cohort size, long-read GS identified potentially high-risk variants in CHD relevant genes.

We also identified a patient where a high-risk non-canonical splice-disrupting variant was only found on short-read GS. We applied a cardiac-specific machine learning model for interpreting canonical and non-canonical splice-disrupting variants developed by us on short-read GS data from patients with cardiac disorders^14^ to both short-read and long-read GS data in this cohort. This identified a heterozygous cryptic splicing variant in a TOF proband in the syndromic CHD gene *PKD1*, which is associated with polycystic kidney disease, but was only called in short-read GS data. Patient-derived myocardial short-read RNA sequencing confirmed variant effect on splicing. The variant was not called in long-read GS data as there were only three reads aligned to the region and a single read supporting the variant (**Figure 9c**). Unfortunately, we were unable to do reverse phenotyping for kidney disease with renal imaging since the patient was only seen at our institution for cardiac interventional procedures, with last follow-up before 10 years of age.

## DISCUSSION

Long-read GS has emerged as an exciting new sequencing technology, providing insights into regions of the genome that have been previously inaccessible or challenging to analyze with short-read GS. However, the genetic etiology of CHD remains elusive in a majority of patients despite the growing use of exome and genome sequencing. The goal of our pilot study was to determine whether long-read GS can address some of the gaps associated with short-read GS by performing a paired comparison of the two sequencing technologies in gene-elusive CHD patients. While a recent study assessed the ability of long-read sequencing to call known pathogenic variants that may be challenging to detect by short-read sequencing^13^, this is, to our knowledge, the first study to directly compare genome-wide and CHD gene variant yield in the same cohort of CHD patients sequenced using both short-read and long-read technologies.

The first important finding was that DNA extracted using conventional protocols can provide good yield of long DNA fragments suitable for long-read GS. This allowed the use of previously processed and stored blood-derived DNA samples in our biobank without requiring prospective blood collection and targeted long fragment DNA extraction protocol.

The second key finding was the significantly higher number of SNVs, deletions, duplications and insertions called with long-read GS, while still having a high agreement with variants called using short-read GS, as previously reported by the All of Us project^19^. For indels, we observed a significantly lower number called in our long-read GS samples, in contrast to previous reports ^20,33^. This discrepancy may be attributed to differences in the computational pipelines used for alignment and variant calling, as these previous studies utilized bwa-mem2 and Genome Analysis Toolkit (GATK) whereas we used Illumina’s DRAGEN pipeline, which has been shown to have better precision and recall for calling indels^34^. However, the low Jaccard index, alongside our observation that most indels were called in low-complexity regions of the genome, suggests that the indels called only in our short-read GS samples were more likely to be false positives^19,20^. In addition, a higher number of deletions, duplications, and insertions were called with long-read GS, which was consistent with previous reports which found that long-read GS was better able to detect these SVs^19,23^. In contrast, a smaller number of inversions were called with long-read GS, which was again consistent with previous findings of inversions being overcalled by short-read GS due to false-positives^19,23^. At a gene level, long-read GS provided comparable coverage of the 99 Tier 1 CHD genes and 48 cardiac relevant genes that reside in so-called ‘dark’ regions of the genome^6^ despite overall lower targeted depth of sequencing^26^.

While overall detection of tandem repeat size was similar (86% of regions) with both sequencing methodologies, for some regions, including in *DAB1*, *BEAN1* and *RFC1*, the larger allele was only detectable using long-read sequencing. Moreover, for other genes, including *HRAS* and *XYLT1*, short-read GS failed to accurately size both the smaller and larger alleles, likely due to the motif length and/or number of repeats, even on the smaller allele. Overall, long-read sequencing enabled the detection and accurate sizing of repeat expansions, including those below the detection limit of short-read GS.

The third key finding was that long-read GS may help in identification of variants in CHD-relevant disease genes that may be missed due to inherent technical limitations of traditional short-read GS. *CFC1, HYDIN* and *NOTCH2* were notable examples of cardiac relevant genes from our study where long-read GS demonstrated more consistent coverage compared with short-read GS for intragenic regions that had low mappability or overlapped with segmental duplications (**Figure 7**, **Table 7**). Of note, intron 10 of *HYDIN* and introns 1 and 4 of *NOTCH2* were intragenic regions where we found a low agreement in variant calls between the two technologies, which reflected lower short-read sequencing depth in these regions. Therefore, it is likely that variants in subregions of these and other cardiac-relevant genes have been underreported in the past, making them excellent candidates for further study in CHD and clinical long-read GS.

Of note, some ‘dark’ genes remained challenging to sequence even with long-read GS. *CFC1B* is 85% ‘dark’ with short-read GS versus 98% ‘dark’ with PacBio long-read GS^6^. Even though our long-read GS data provided relatively better coverage of *CFC1B* than short-read GS data, the sequencing depth was still too low to make confident variant calls with long-read GS. Oxford Nanopore is currently the only reported long-read technology that can 100% resolve *CFC1B*^35^.

*SMN1* is another example of a gene that has been historically challenging to sequence due to its high sequence similarity with its paralog *SMN2*. 94.6% of the coding sequence of *SMN1* is ‘dark’ on short-read GS compared to only 2% of the coding sequence being ‘dark’ on long-read GS^6^. In our cohort, *SMN1* had better coverage with long-read GS but intragenic regions such as exon 1 and intron 1 remained poorly covered by both long-read and short-read with an average sequencing depth of less than 10×. Ebbert et al.^6^ also demonstrated that PacBio did not perform as well as other long-read sequencing platforms such as Oxford Nanopore Technologies and 10x Genomics for sequencing the *SMN1* gene^35^. Other cardiac relevant genes of note are *H19* and *KCNE1*. In our cohort, short-read GS failed to sequence the *H19* gene, whereas long-read GS performed only slightly better with a sequencing depth of under 10× for most samples. Both long-read and short-read GS struggled to sequence the intragenic region of *KCNE1* encompassing exon 3, intron 3 and exon 4. The difficulty in sequencing these genes may be explained by the presence of segmental duplications and low mappability^27^.

Long-read GS did not identify any definitively pathogenic variants in Tier 1 CHD genes in this relatively small genotype-elusive cohort, with determination of *de novo* variant inheritance, which account for a majority of sporadic CHD^3^, being difficult in the absence of parental long-read GS. Nonetheless, it did allow us to resolve additional variants that may contribute to CHD and that are not typically captured by short-read sequencing based clinical genetic tests. Further, our study represents one of the first attempts at resolving ‘dark’ regions of CHD genes. While variants were identified in these ‘dark’ regions, it was not possible to determine their pathogenicity since they have not been captured in previous short-read sequencing studies.

While there are technical advantages to long-read GS technology, there are also differences in cost, with a ∼30× short-read genome costing less than half of the ∼20× long-read genome at the time of our data production. For short-read GS, we aimed for a 30× genome-wide sequencing depth. This was shown nearly two decades ago to be accurate and sufficient for most genomic applications^36^, and has since been used as the standard sequencing depth for major genome sequencing projects^37,38^. For long-read GS, we aimed for a 20× genome-wide sequencing depth as this was shown to provide good precision and recall of variants, with the sensitivity for detecting true positive SNVs and SVs in particular nearing 1 by this depth^39^. A lower but acceptable genome-wide sequencing depth in the long-read data was enabled both by the longer read length and also by the higher accuracy of those reads. It is important to note, however, that as both short-read and long-read sequencing technologies continue to improve in their technical specifications, computational workflows, and cost, future projects should utilize the platform, data production, and workflows that best suit those needs.

Overall, the strength of our study was the paired comparison of genomic data using short and long-read technologies which helped identify areas where long-read GS has an advantage over short-read GS, but also identified the limitations of long-read GS overall, when applied to CHD patients. Nonetheless, our findings suggest that there may be a role for applying long-read GS to identify putatively pathogenic variants in patients who are otherwise genotype-elusive. However, this requires a larger study of CHD patients and family members, as well as a cost-effectiveness analysis to determine whether the increased diagnostic yield justifies the higher cost of long-read sequencing.

## METHODS

### Study cohort

The long-read GS study cohort included 43 unrelated probands with CHD, of which 36 had TOF and 7 had TGA **(Table 1)**. All individuals were enrolled through the Heart Centre Biobank Registry at the Hospital for Sick Children (Toronto, Ontario, Canada)^40^. Collection and use of biospecimens through the registries was approved by local Research Ethics Boards and written informed consent was obtained from all patients and/or their parents/legal guardians and study protocols adhered to the Declaration of Helsinki. Sample identifiers used in this study were not known to anyone outside the research group.

### DNA extraction and quantification

2–4 ml blood was collected in EDTA tubes and DNA used for GS was extracted from blood through chemagic™ STAR robotic system using a magnetic bead methodology at The Centre for Applied Genomics (TCAG, The Hospital for Sick Children) and stored at –20 to 4 degrees C in the biobank. An additional fresh blood sample from a biobank enrolled patient was used for long fragment DNA extraction using a targeted protocol to compare long fragment yield between the two extraction methods. The NEB Monarch High Molecular Weight DNA extraction kit for ‘fresh nucleated blood’ protocol was used (New England Biolabs, Ipswich, MA, Cat#T3050L). The highest recommended setting of 2000 RPM was used for the lysis step and 125 μl of elution buffer was used.

DNA was quantified using the Qubit dsDNA High Sensitivity Assay and sample purity was checked using Agarose Gel Electrophoresis and NanoDrop OD 260/280 ratio. DNA samples were deemed to have met quality control thresholds if they had a single clear band on the agarose gel and a 260/280 absorbance ratio greater than 1.3. DNA sample quality was further assessed on the Genomic DNA ScreenTape on the Agilent TapeStation (TCAG, The Hospital for Sick Children) to calculate DNA integrity number (DIN) and fragment sizes. DIN determines the fragmentation of a genomic DNA sample by assessing the distribution of signal across the size range, and applies an automatically calculated number (range 1-10). A DIN ≥7 indicates highly intact genomic DNA, and a low DIN indicates a degraded genomic DNA sample. 5-7 ug of DNA was sheared using Covaris g-TUBEs to 15-18 kb and rechecked on the TapeStation to confirm the fragment size before library prep. Sheared DNA was used as input for library preparation using the SMRTbell Prep Kit 3.0 following the manufacturer’s recommended protocol. DNA was end-repaired, ligated with SMRTbell barcoded adapters, and size-selected using either Ampure PB bead size-selection or the BluePippin System to remove fragments below 10 kb. Libraries were validated on the TapeStation to check for size and quantified by the Qubit 3.0 Fluorometer ds DNA High Sensitivity Assay.

Analyses and plotting were performed using R v4.4.1. The significance of correlations were assessed using the ‘cor.test()’ function with Pearson correlation. Plotting used the ‘ggplot2’ library with the ‘ggpattern’ library for striped coloring.

### Short-read genome sequencing and data processing workflows

Short-read GS was performed on high quality DNA from blood or saliva using the Illumina HiSeq X or NovaSeq platform (TCAG, The Hospital for Sick Children, Toronto, Canada; Psomagen, Maryland, USA). Illumina TruSeq DNA PCR-Free kits were used for library preparation.

FASTQ files were processed using DRAGEN Bio-IT Platform v3.8.4 (https://support-docs.illumina.com/SW/DRAGEN_v38/Content/SW/DRAGEN/GPipelineIntro_fDG.htm). Briefly, the reads were aligned to GRCh38 human genome reference (hg38-alt-aware-graph). Small variants (SNV and indel), CNVs, SVs, HLA and tandem repeats were called. Files in standard output format were subsequently generated – CRAM for alignment and VCF for small variants, CNVs and SVs. SV calling was performed by Manta through the DRAGEN pipeline and CNV calling by the DRAGEN CNV caller. SV and CNV calling were performed using default parameters, with the exception that the CNV threshold was instead set to 5 kb. Per sample analysis metrics were generated. The small variant calls were annotated using an ANNOVAR based pipeline and the CNVs and SVs were annotated using custom scripts available at https://github.com/ccmbioinfo/mital-chd-pacbio.

### Long-read genome sequencing and data processing workflows

DNA extractions considered for long-read GS were required to have at minimum 70% read fragments ≥ 20 kb in size. A total of 5-7 μg of DNA at a minimum concentration of 20 ng/μl was used for long-read GS. Validated libraries were set up for sequencing on the PacBio Sequel IIe loading on the 8M SMRTCell for a movie time of 30 hours. Long-read GS was performed at TCAG, The Hospital for Sick Children, Toronto, Canada.

Unaligned BAM files were processed using the PacBio Human WGS Workflow (https://github.com/PacificBiosciences/pb-human-wgs-workflow-snakemake). Briefly, the reads were aligned to GRCh38 human genome reference. Small variants (SNV and indel), structural variants (including CNVs), and tandem repeats were called. Files in standard output format were subsequently generated – BAM for alignment and VCF for small variants, CNVs and SVs. SNV and indel calling was done by DeepVariant, and SV calling by pbsv. Per sample analysis metrics were generated. The small variant calls were annotated using an ANNOVAR based pipeline and the CNVs and SVs were annotated using custom scripts available at https://github.com/ccmbioinfo/mital-chd-pacbio..

### Sequencing data quality control

Mosdepth v0.3.4 was used to calculate the average sequencing depth across long-read and short-read genomes^41^. Somalier v0.2.13 was used to estimate sex, relatedness and ancestry on all genomes^42^. The predictions were manually compared to the self-reported sex, ancestry, and relationships to check for any sample swaps or unexpected relationships. Plots were generated using the ggplot2 library (https://ggplot2.tidyverse.org) and patchwork library (https://patchwork.data-imaginist.com/index.html) to panel the plots together.

### CHD gene lists

Tier 1 CHD genes included genes with moderate, strong, or definitive association with CHD according to the ClinGen criteria^15^. This gene list consisted of 17 isolated CHD and 82 syndrome-associated CHD genes yielding a total of 99 genes. This gene list served as our primary gene list for all analyses detailed in this paper.

For the sequencing depth analysis, we expanded our primary Tier 1 CHD gene list to also include cardiac relevant genes that reside in the ‘dark’ regions of the genome. The Twist Alliance Dark Genes panel is a publicly available resource consisting of 389 genes that are difficult to sequence with short-read GS^26^. It is available to download as a BED file for research purposes from their website (https://www.twistbioscience.com/resources/data-files/twist-alliance-dark-genes-panel-bed-file). To extract cardiac relevant genes from this dark gene panel, we intersected our in-house CHD gene list (consisting of 1100 genes) with the Twist panel using bedtools. This yielded a total of 56 genes which included 8 Tier 1 CHD genes (*ADAMTS10*, *B3GAT3*, *BRAF*, *EHMT1*, *FLT4*, *G6PC3*, *KANSL1*, and *PKD1*) that were already a part of our primary gene list for analysis and 48 additional cardiac relevant genes that we included for the sequencing depth analysis.

Gene information was annotated using Online Mendelian Inheritance in Man (OMIM)^43^ and Clinical Genome Resource (ClinGen)^44^ **(Table 3)**. Canonical transcriptional isoforms were annotated using Matched Annotation from NCBI and EMBL-EBI (MANE)^45^. Gene constraint annotations were obtained from the Genome Aggregation Database (gnomAD) (v4.1) (https://gnomad.broadinstitute.org/news/2024-03-gnomad-v4-0-gene-constraint/).

### Comparing variant calls

*Small Variants:* VCFs for both short-read and long-read were decomposed and normalized using vt v0.5 followed by filtering for PASS variants in autosomes, chrX, chrY, and chrM^46^. To determine the total number of SNVs and indels genome-wide, the‘bcftools view’ command was utilized with the‘-H’ (no header) and‘-v’ (variant types, either SNVs or indels) parameters, followed by the‘wc –l’ command for each sample. To retrieve the total number of SNVs and indels that intersect with Tier 1 CHD genes, we first downloaded BED files with the coordinates for Tier 1 CHD genes from UCSC Table Browser using the following specifications: Clade: Mammal, Genome: Human, Assembly: Dec. 2013 (GRCh38/hg38), Group: Genes and Gene Predictions, Track: NCBI RefSeq, Table: RefSeq Select and MANE (ncbiRefSeqSelect), Region: Genome, Identifiers: List of Tier 1 CHD Genes, Output format: BED – browser extensible data. Next, the‘bcftools view’ command was utilized with the‘-H’, ‘-v’ and an additional‘-R’ parameter (regions file downloaded from UCSC), followed by the‘wc –l’ command for each sample. In R, we performed a paired two-sided Wilcoxon test to evaluate whether there was a significant difference in the number of SNVs and indels called in the two technologies (long-read GS and short-read GS). This testing was performed for genome-wide variant calls as well as variants that intersect with CHD Tier 1 genes. Additionally,‘bcftools isec’ was used to calculate the number of SNVs and indels unique to each technology and variants shared between the two technologies. We next calculated the Jaccard index as the number of variants that intersect between long-read and short-read divided by the union of variants called in short-read and long-read. The Jaccard indices were calculated for all sample pairs both genome-wide and in CHD Tier 1 genes. The average of the values for all participants were reported in the results. For SNVs, fold changes were calculated by taking the average ratio of long-read to short-read SNVs called across patients. For indels, fold changes were calculated by taking the average ratio of short-read to long-read indels called across patients. Plots were generated using the ggplot2 library (https://ggplot2.tidyverse.org) and patchwork library (https://patchwork.data-imaginist.com/index.html) to panel the plots together.

*CNV/SV:* Both long-read and short-read SV/CNV ‘.vcf’ files were parsed into ‘.tsv’ reports. Using the R package dplyr (https://dplyr.tidyverse.org), the reports were filtered down to include only PASS variants, and for CHD-specific analyses, only variants annotated to overlap with a Tier 1 CHD gene were retained. Further, an additional size filter was added to remove long-read variants less than 50 bp long. Variant counts were attained post-filtering using the table() function in R to tabulate the number of variants per variant type. To compare structural variants called genome-wide and in Tier 1 CHD genes, we applied a ‘PASS’ filter on all variants, and an additional 50 bp + filter on SVs called by pbsv in the long-read workflow to facilitate comparisons with short-read, as short-read SV callers by default only call variants > 50 bp.

To investigate how well SV calls matched between long-read and short-read, long-read SVs were intersected with short-read SVs called by Manta, using bedtools intersect. A long-read SV was considered matching if had a 50% reciprocal overlap with a short-read SV, which was determined by setting the –f and –F flags of the bedtools intersect command to 0.5. The Jaccard index was determined by dividing the number of long-read variants that had a short-read match, by the total number of variants called in both long-read and short-read. Fold changes were calculated by taking the average ratio of long-read to short-read variants called across patients, for each variant class. Concordance was calculated as the number of long-read insertions with short-read matches, divided by the total number of long-read insertions.

### Tandem repeat expansion workflow

Within each region and for each sample, we defined the smaller and larger allele, then calculated Pearson’s correlation coefficients and p-values between the long-read sequencing data and short-read sequencing data for the smaller and larger allele size separately. Next, we used correlation coefficients on a smaller allele and correlation coefficient on a larger allele (**Table 5**) to compare the performance of repeat sizing tools across the 59 regions.

We used the coordinates and repeat motif sequence in 59 known disease-causing tandem repeat regions identified in 18 previous studies (**Table 5**) to determine repeat size using ExpansionHunter^25^ in all 43 short-read GS samples. For each region and sample, we designated a ‘smaller’ allele, and a ‘larger’ allele based on their relative sizes. Similarly, we used the Tandem Repeat Genotyping Tool^24^ to analyze 43 long-read GS samples in the same 59 known disease-causing tandem repeat regions (**Table 5)**.

Within each region, we calculated the genotype frequency by counting the number of instances of a smaller allele and larger allele combination among the patients, separately for long-read data and short-read data. Further, we analyzed the distribution of repeat size on the smaller (or larger) allele in long-read sequencing data versus the distribution of repeat size on the smaller (or larger) allele in short-read sequencing data. This allowed us to calculate Pearson’s correlation coefficient and corresponding p-value. The correlation coefficients and p-values for both smaller and for larger allele across all 59 regions are listed in **Table 5**. If the repeat was not identified in certain samples, potentially due to issues such as low quality or poor coverage of the region, we restricted our analysis to samples for which data were available for this region in both datasets when generating the plot and calculating the correlation. Notably, in the final plot, we excluded the repeat near *ZNF713* due to a limited number of samples (n = 7) where the repeat was detected.

### Sequencing depth analysis for CHD genes

Regions of interest for the sequencing depth analysis included (i) Tier 1 CHD genes and (ii) cardiac relevant genes on the Twist panel (**Table 3**). Mosdepth v0.3.4 was used to calculate the overall sample sequencing depth as well as the sequencing depth of the regions of interest in all paired long-read and short-read samples. To assess the differences in sequencing depth, we calculated the normalized sequencing depth for each gene of interest per sample by dividing the gene sequencing depth by the sample genome-wide sequencing depth. For each gene, a two-sided Wilcoxon test was performed on the normalized sequencing depths in all samples, followed by p-value correction using the Benjamini-Hochberg procedure to account for multiple testing. Genes with a FDR <0.05 were considered to have a significant difference in depth. For such genes, the median normalized sequencing depths were further investigated to determine which intragenic regions had a higher depth in long-read versus short-read GS.

To compare the exonic and intronic sequencing depth differences for *CFC1, HYDIN, NOTCH2*, *CFC1B*, *H19*, *KCNE1* and *SMN1*, we first downloaded BED files containing coordinates of exons and introns from UCSC table browser and then followed the same method as described above to short-list regions that are differentially covered in long-read and short-read GS. To compare variant calls, we used‘bcftools isec’ to calculate the number of SNVs and indels unique to each technology and variants shared between the two technologies followed by calculating the Jaccard index for each gene sub-region per sample. The median of the Jaccard indices were reported in the results.

Plots were generated using the ggplot2 library (https://ggplot2.tidyverse.org) and patchwork library (https://patchwork.data-imaginist.com/index.html) to panel the plots together.

### Identification of putatively pathogenic and high-risk variants

Variant analyses were performed for each patient, for both their short-read and long-read genomes, across the list of attached CHD genes, as described in our previous study^14^ and summarized below. Interpretation was performed in line with the American College of Medical Genetics and Genomics and the Association of Molecular Pathology (ACMG/AMP) criteria for sequence-level variants^16^ and constitutive CNVs^17^. Parental data was used whenever available to phase variants or establish them as *de novo*.

#### Short-read genome analysis

For sequence-level variants, annotation of short-read genomes was performed using InterVar^47^ v2.0.2 20190327. The Human Gene Mutation Database (HGMD) Pro 2019 database was used through InterVar to assign a PS3 (strong evidence of pathogenicity) criterion if a classification of disease-associated polymorphism with supporting functional evidence was in the database, or PP5 (supporting evidence of pathogenicity) if the variant in question was annotated as a disease-associated polymorphism (DFP) or disease-causing mutation (DM). Following annotation by InterVar, variants designated as “Pathogenic” or “Likely Pathogenic” were manually reviewed. Variants were queried through ClinVar to identify other reports of pathogenicity, or other variants in the same position. Population allele frequencies were attained from gnomAD v3.1.2, while gene constraint metrics were retrieved from gnomAD v2.1.1. Variant quality was checked both by examining regional mappability, through the UCSC Genome Browser RepeatMasker track, and also by validating reads supporting the variant in question using the Integrative Genome Browser. Heterozygous variants that had an allele fraction less than 33% or more than 66% were excluded, as were those with less than 20× depth, and those with several base mismatches in neighboring reads, in order to reduce the rate of false positives.

For SVs and CNVs, variants were filtered out if they were present at greater than 1% frequency in an internal database generated from Illumina HiSeq X sequencing data at TCAG^48^. Variants were further filtered out if they did not overlap with an exonic region of a CHD gene or established ISCA region associated with CHD. Only ‘PASS’ variants were considered. Variants were then searched through the Decipher browser, and those that overlapped substantially with population variants in the Database of Genomic Variants (DGV) or gnomAD Structural Variants tracks were not further considered. Called inversions were excluded from further analysis if one or both breakpoints were not located within a CHD gene. Remaining variants were validated visually using Samplot or the Integrative Genomics Viewer (IGV), and then manually classified using the ACMG/AMP criteria for constitutive CNVs. Deletions and duplications with breakpoints in a single gene were assessed for the PVS1 (very strong indicator of pathogenicity) criterion using AutoPVS1.

#### Long-read genome analysis of sequence-level variants

Sequence level variants were further annotated with variant allele frequencies from the Genomic Answers for Kids (GA4K) PacBio cohort (n=960)^31^ and dark region from the Twist panel^26^.

Variants that had homozygotes in gnomAD v3.0, greater than 50 alleles in gnomAD v.3.0, greater than 100 alleles in GA4K cohort and variants that did not overlap with CHD genes were filtered out. Variants in genes with phenotypic association were analyzed according to the ACMG guidelines. Inheritance of these variants were determined for probands that had parental short-read data available through visual inspection in IGV when possible.

*Putatively splice-disrupting SNVs and indels:* Both short-read and long-read variants were identified using the previously described workflow^14^. Pre-computed masked SpliceAI^49^ delta scores were utilized where possible (https://basespace.illumina.com/projects/66029966), otherwise SpliceAI (v1.3.1) was used to generate masked delta scores with a maximum distance of 100bp between the variant and gained/lost splice site. Exome annotations and splice junctions were similarly obtained from SpliceAI. SNVs and indels with a ‘PASS’ flag were extracted using bcftools v1.9^50^. Variants with a SpliceAI delta score ≥ 0.2 were retained and subsequently annotated with the predicted effect (VEP v102^51^), reported pathogenicity (ClinVar 2022-04-03 and HGMD Pro 2019), control allele frequency (gnomAD v2.1.1 and v3.1.2), gene constraint (gnomAD v2.1.1), genomic low complexity regions (https://github.com/lh3), genomic RepeatMasker regions (https://www.repeatmasker.org), and wild-type splicing branchpoints^52,53^. Ensembl RNA transcripts were further annotated as canonical by MANE v1.0 (MANE Select or MANE Plus Clinical)^45^. An optimal weighted cardiac-specific machine learning model was then used to select for high-confidence splice-disrupting variants^14^. Variants were further filtered to limit to those with a gnomAD PopMax filtered allele frequency < 0.0001 and absent from the GA4K cohort. These filtered, high-confidence variants were then analyzed in corresponding myocardial short-read RNA sequencing data from the same patient to determine if they were associated with splicing events.

SVs/CNVs were further annotated using variant allele frequencies from the GA4K PacBio cohort^31^, using 50% reciprocal overlap, and variants present in the GA4K database at a frequency greater than 1% were filtered out, as were variants less than 50 bp in length, and those that did not overlap with exonic regions of CHD genes. Variants were further filtered after Decipher and UCSC genome browser queries as described above and classified using the ACMG/AMP criteria for constitutive CNVs and AutoPVS1.

### Statistical Analyses

A Pearson correlation test was used to assess the significance of the association between input DNA fragment sizes or DIN and mean long-read sequenced read lengths.

To evaluate whether there was a significant difference in the frequency of SNVs and indels called by the two technologies (long-read GS and short-read GS), we conducted a paired two-sided Wilcoxon test. This was done for genome-wide variant calls as well as variant calls intersecting CHD tier 1 genes. Results with a nominal p-value < 0.05 were considered to be significant. Furthermore, results from GATK concordance v4.0.0 were used to calculate the Jaccard indices for all sample pairs both genome-wide and in CHD Tier 1 genes. The average of the values for all participants were reported in the results.

A Pearson correlation test was used to assess the significance of the association between short-read and long-read tandem repeat allele size estimates.

For the sequencing depth analysis of CHD genes, we first calculated the normalized sequencing depth for each gene per sample by dividing the gene sequencing depth by the sample genome-wide sequencing depth. Next, we performed a two-sided Wilcoxon test on the normalized sequencing depths of all samples, followed by p-value correction using Benjamini-Hochberg procedure to account for multiple testing. Genes with an FDR < 0.05 were considered to have a significant difference in the normalized sequencing depth. For genes with a significant difference, the differences in medians of normalized sequencing depths were further compared manually to determine genes that had a higher normalized median sequencing depth in long-read versus short-read GS. The same analysis methodology was followed to determine significant differences in normalized sequencing depth of intergenic regions of *CFC1, HYDIN, NOTCH2*, *CFC1B*, *H19*, *KCNE1* and *SMN1*.

To evaluate differences in size for SV classes, a two-sided Wilcoxon test was performed. All statistical tests were performed using the R programming language.

## ETHICS

The institutional Research Ethics Board of The Hospital for Sick Children, Amsterdam Medical Center, The Children’s Hospital at Westmead, and other participating sites gave ethics approval for the collection and use of data and biospecimens from patients enrolled through respective registries The Heart Centre Biobank (Ontario, Canada), CONCOR (Amsterdam, Netherlands), and Kids Heart BioBank (Sydney, Australia). The study protocols adhered to the Declaration of Helsinki. Written informed consent was obtained from all patients and/or their parents/legal guardians.

## DATA AVAILABILITY

Genomic data for short-read GS samples are available in the European Genome-Phenome Archive (EGA) under accession EGAS50000000586 (https://ega-archive.org/studies/EGAS50000000586), and will be available for download upon approval by the Data Access Committee (https://theheartcentrebiobank.com/sample-request). Sequencing data for the long-read GS samples are being deposited in the Canadian Genome-Phenome Archive, part of the Federated EGA. The GitHub library with the code for calculation and visualization of tandem repeat size distribution in long-read versus short-read data is available on Zenodo under “TR comparison in LONG vs SHORT read data” DOI: “10.5281/zenodo.10637193”.

## AUTHORS CONTRIBUTIONS

RL and SM conceived and designed the work. RL, AJ, NH, AM, and SM drafted the work and substantively revised it. RL, AJ, NH, AM, and YY performed bioinformatics and statistical analyses. SM and RKCY acquired funding for the project. All authors contributed to acquisition, analysis or interpretation of data. All authors read and approved the final manuscript, and have agreed both to be personally accountable for the author’s own contributions and to ensure that questions related to the accuracy or integrity of any part of the work, even ones in which the author was not personally involved, are appropriately investigated, resolved, and the resolution documented in the literature.

## ACKNOWLEDGEMENTS

We thank The Centre for Applied Genomics at the Hospital for Sick Children for performing genome sequencing. We thank Madeline Couse for setting up long-read annotation workflows and aiding in variant interpretation. We also thank the patients and family members from the Heart Centre Biobank who participated in this study.

This study makes use of data generated by the DECIPHER community. A full list of centres who contributed to the generation of the data is available from https://deciphergenomics.org/about/stats and via email from. DECIPHER is hosted by EMBL-EBI and funding for the DECIPHER project was provided by the Wellcome Trust [grant number WT223718/Z/21/Z].

