## Supplemental Figures for "Long-read genome sequencing increases genomic yield in congenital heart disease"

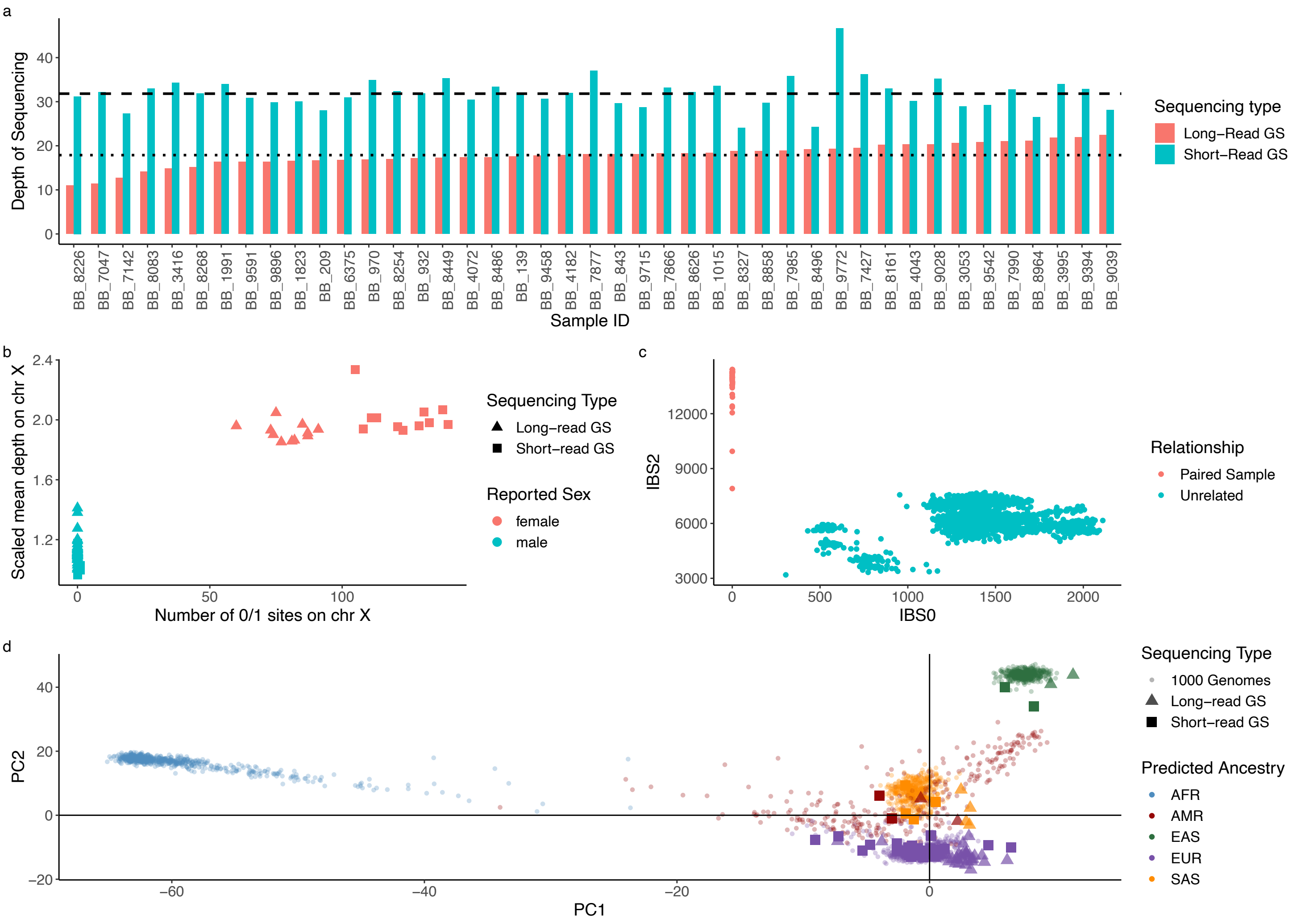


**Supplemental Figure S1. Sequencing depth and quality control metrics in long-read and short-read GS. (a)** Sequencing depth: The average depth of sequencing for the 43 CHD proband samples in long-read GS (red bars) was 17.82× (dotted line) and in short-read GS (blue bars) was 31.88× (dashed line). **(b)** Sex QC: scaled mean depth on chromosome X on the y-axis versus the number of heterozygous sites on chromosome X on the x-axis, showing paired long-read (triangles) and short-read (squares) GS of 43 CHD proband samples. Males (blue) and females (red) clustered separately on the plot with no sex mismatches. **(c)** Sample to sample relatedness: A comparison of the IBS0 (number of sites where 1 sample is homozygous reference and another is homozygous alternate) and IBS2 (number of sites where samples have the same genotype) metric for the 43 CHD probands. Each point is a pair of samples. All long-read and short-read sample pairs (red) have an IBS0 close to zero and a higher IBS2 and vice versa for the unrelated sample pairs (in blue). **(d)** Cohort Ancestry prediction. This plot depicts the ancestry prediction of the 43 CHD probands using long-read GS (triangles) and short-read (squares) overlaid on 1000 Genomes Project data (small circles). Ancestry prediction was concordant between both sequencing technologies. CHD, congenital heart disease; GS, Genome sequencing; IBS, Identity-by-state; QC, Quality control

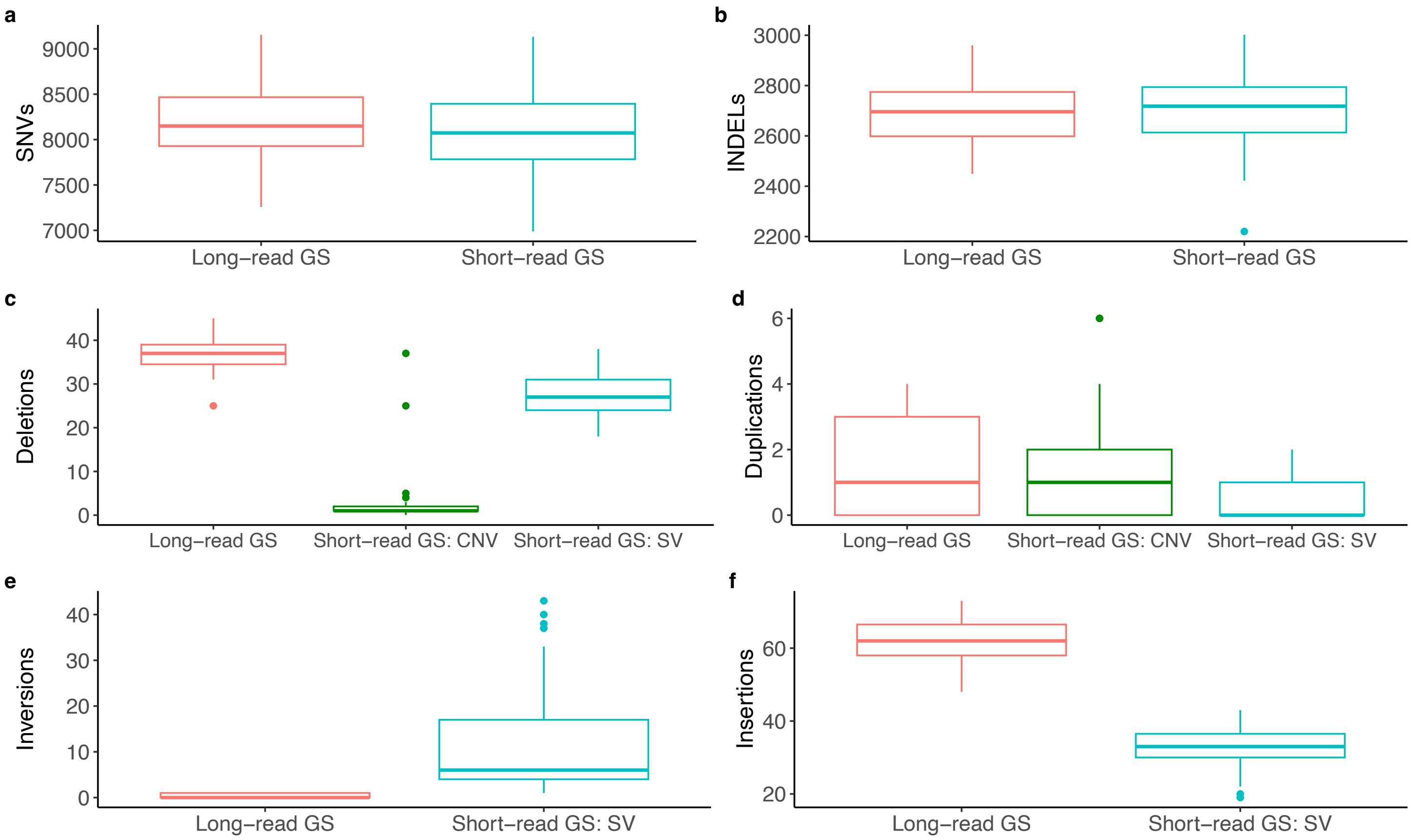


**Supplemental Figure S2. CHD gene variant yield in long-read and short-read genome sequencing**. Box plots comparing variants called in long-read and short-read GS that overlap 99 Tier 1 CHD genes **(a)** SNVs. **(b)** Indels. **(c)** Deletions. **(d)** Duplications. **(e)** Inversions. **(f)** Insertions. In CHD genes, long-read GS called more SNVs (1.01-fold; nominal p-value = $4.13\times10^{-5}$), deletions (1.39-fold; nominal p-value = $5.2\times10^{-23}$), duplications (nominal p-value = $5.8\times10^{-5}$, fold-change unavailable due to low per-sample duplication numbers), and insertions (1.94-fold; nominal p-value = $1.4\times10^{-15}$) than short-read GS but fewer indels (0.999-fold; nominal p value = $1.37\times10^{-3}$) and inversions (0.035-fold; nominal p-value = $3.9\times10^{-16}$). These trends were comparable with genome-wide variant yield. Short-read GS: CNV designates structural variants called by copy number variant callers, while Short-read GS: SV designates structural variants called by structural variant callers. Asterisk denotes significant differences (nominal p-value $< 0.05$).
CHD, congenital heart disease; SNV, single nucleotide variants; Indels, small insertion-deletions; CNV, copy number variants; SV, structural variants

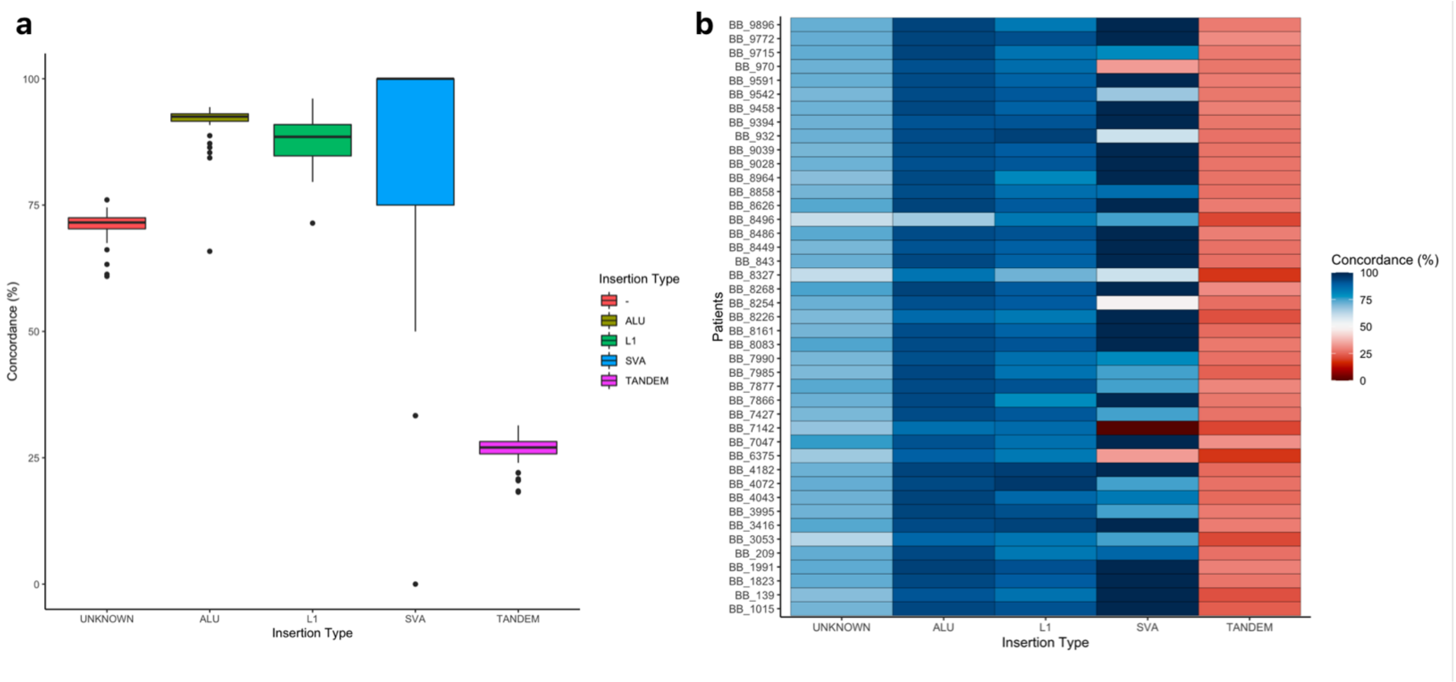


**Supplemental Figure S3: Concordant insertion calls between long-read and short-read genome sequencing.** Concordance was calculated as the percentage of long-read insertions that were also called in short-read data, stratified by insertion type, (**a**) at the cohort level, and (**b**) in individual participants. While there was a high agreement between long-read GS and short-read GS in detecting most insertion types, there was a much lower concordance for tandem repeat insertion calls.

SV, structural variant; GS, genome sequencing; INV, inversion; DEL, deletion; INS, insertion; DUP, duplication; ALU, Alu element; L1, long interspersed nuclear element-1; SVA, SINE-VNTR-Alu retrotransposon

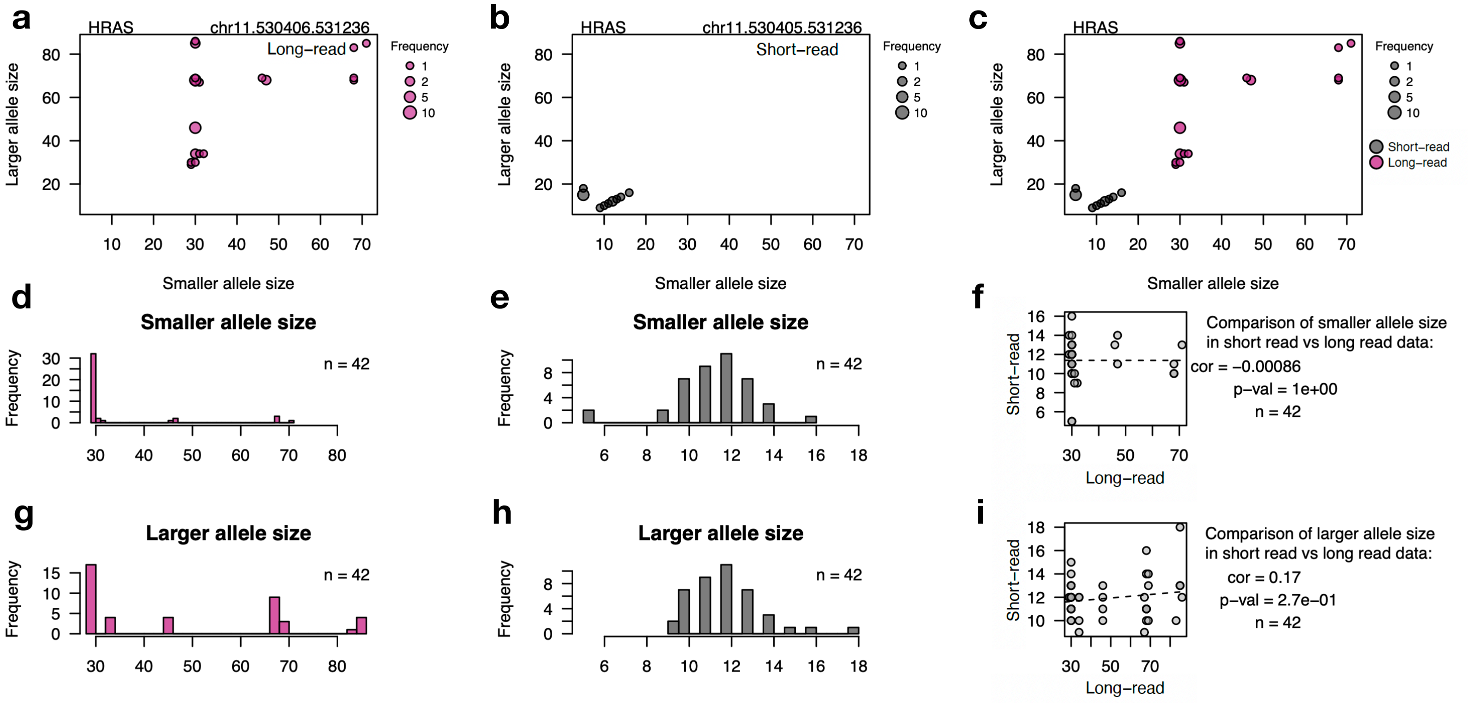


**Supplemental Figure S4. *HRAS* tandem repeat sizing using short-read and long-read sequencing.** GGCGTCCCCTGGAGAGAAGGGCGAGTGT-repeat near *HRAS* gene detected using **(a)** long-read sequencing (pink), **(b)** short-read sequencing (grey), and **(c)** comparison of the two approaches; x-axis represents repeat size on a smaller allele, y-axis represents repeat size on a larger allele, each dot represents a genotype, the size of the dot represents number of samples with the genotype. Smaller and larger allele size estimates did not overlap across platforms, indicating broadly different repeat size estimates. Distribution of repeat size on a smaller allele using **(d)** long-read and **(e)** short-read sequencing. **(f)** Comparison of a smaller allele size in short vs long-read sequencing data. Distribution of repeat size on a smaller allele using **(g)** long-read and **(h)** short-read sequencing. **(i)** Comparison of a larger allele size in short versus long-read sequencing data. Estimates were not significantly correlated for either the smaller or larger allele between the two platforms, indicating broad disagreement in estimates of repeat size.

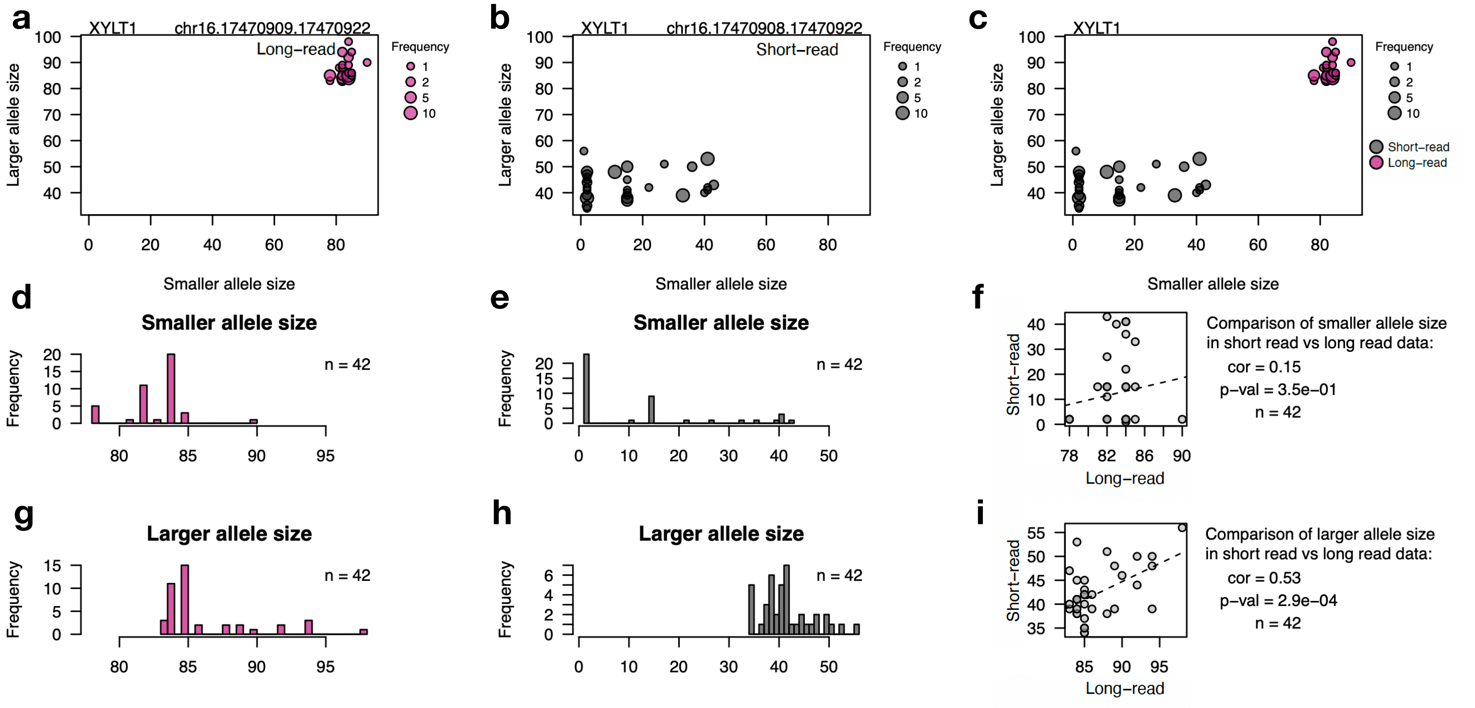


**Supplemental Figure S5.** ***XYLT1* tandem repeat sizing using short-read and long-read sequencing.** CGG-repeat near *XYLT1* gene detected using **(a)** long-read sequencing (pink), **(b)** short-read sequencing (grey), and **(c)** comparison of the two approaches; x-axis represents repeat size on a smaller allele, y-axis represents repeat size on a larger allele, each dot represents a genotype, the size of the dot represents number of samples with the genotype. Smaller and larger allele size estimates did not overlap across platforms, indicating broadly different repeat size estimates. Distribution of repeat size on a smaller allele using **(d)** long-read and **(e)** short-read sequencing. **(f)** Comparison of a smaller allele size in short vs long-read sequencing data. Distribution of repeat size on a smaller allele using **(g)** long-read and **(h)** short-read sequencing. **(i)** Comparison of a larger allele size in short versus long-read sequencing data. The estimates for the smaller alleles were not significantly correlated between the two platforms. Although estimates for the larger allele were significantly correlated, the scale of estimates did not overlap, indicating similar trends despite broadly different absolute repeat size estimates.

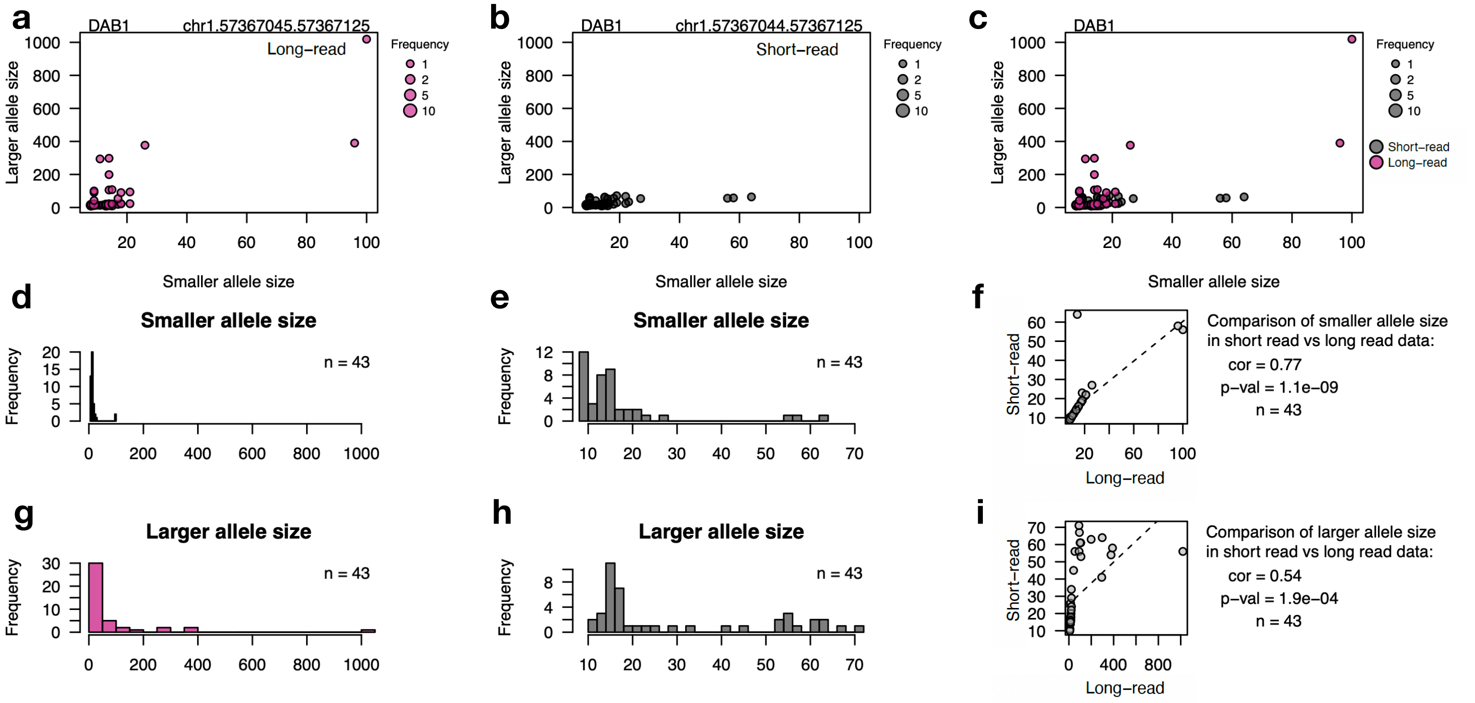


**Supplemental Figure S6.** ***DAB1* tandem repeat sizing using short-read and long-read sequencing.** AAAAT-repeat near *DAB1* gene detected using **(a)** long-read sequencing (pink), **(b)** short-read sequencing (grey), and **(c)** comparison of the two approaches; x-axis represents repeat size on a smaller allele, y-axis represents repeat size on a larger allele, each dot represents a genotype, the size of the dot represents number of samples with the genotype. Smaller and larger allele size estimates are similar for most participants across either platform, but only long-read sequencing detected large outlier allele sizes in participants. Distribution of repeat size on a smaller allele using **(d)** long-read and **(e)** short-read sequencing. **(f)** Comparison of a smaller allele size in short vs long-read sequencing data. Distribution of repeat size on a smaller allele using **(g)** long-read and **(h)** short-read sequencing. **(i)** Comparison of a larger allele size in short versus long-read sequencing data. While the estimates for both smaller and larger allele were significantly correlated between the two platforms, the estimates of the smaller allele were more highly correlated, indicating that long-read sequencing provided more accurate estimates for the larger allele.
